# South African Population Immunity and Severe Covid-19 with Omicron Variant

**DOI:** 10.1101/2021.12.20.21268096

**Authors:** Shabir A. Madhi, Gaurav Kwatra, Jonathan E. Myers, Waasila Jassat, Nisha Dhar, Christian K. Mukendi, Amit J. Nana, Lucille Blumberg, Richard Welch, Nicoletta Ngorima-Mabhena, Portia C. Mutevedzi

## Abstract

**Background:** We conducted a seroepidemiological survey from October 22 to December 9, 2021, in Gauteng Province, South Africa, to determine SARS-CoV-2 immunoglobulin G (IgG) seroprevalence primarily before the fourth wave of coronavirus disease 2019 (Covid-19), in which the B.1.1.529 (Omicron) variant was dominant. We evaluated epidemiological trends in case rates and rates of severe disease through to January 12, 2022, in Gauteng.

**Methods:** We contacted households from a previous seroepidemiological survey conducted from November 2020 to January 2021, plus an additional 10% of households using the same sampling framework. Dry blood spot samples were tested for anti-spike and anti-nucleocapsid protein IgG using quantitative assays on the Luminex platform. Daily case, hospital admission, and reported death data, and weekly excess deaths, were plotted over time.

**Results:** Samples were obtained from 7010 individuals, of whom 1319 (18.8%) had received a Covid-19 vaccine. Overall seroprevalence ranged from 56.2% (95% confidence interval [CI], 52.6 to 59.7) in children aged <12 years to 79.7% (95% CI, 77.6 to 81.5) in individuals aged >50 years. Seropositivity was more likely in vaccinated (93.1%) vs unvaccinated (68.4%) individuals. Epidemiological data showed SARS-CoV-2 infection rates increased and subsequently declined more rapidly than in previous waves. Infection rates were decoupled from Covid-19 hospitalizations, recorded deaths, and excess deaths relative to the previous three waves.

**Conclusions:** Widespread underlying SARS-CoV-2 seropositivity was observed in Gauteng Province before the Omicron-dominant wave. Epidemiological data showed a decoupling of hospitalization and death rates from infection rate during Omicron circulation.

## BACKGROUND

The B.1.1.529 (Omicron) variant of severe acute respiratory virus syndrome coronavirus 2 (SARS-CoV-2) was first reported on November 25, 2021, in Gauteng Province, South Africa.^1^ The World Health Organization designated Omicron a variant of concern due to its predicted greater transmissibility and its potential to evade vaccine-induced and natural infection-induced neutralizing antibody immunity.^2^ The Omicron variant contains mutations that indicate it could be more infectious, more transmissible, and possibly better able to evade innate immunity and neutralizing antibody activity compared with the wild-type (WT) variant.^3-5^ In addition to at least 32 mutations affecting the spike protein,^6^ the Omicron variant harbors three mutations affecting the membrane protein and six involving the nucleocapsid protein, compared with only seven spike and one nucleocapsid-protein mutation in the antibody-evasive B.1.351 (Beta) variant.^7^

The Omicron variant out-competed the B.1.617.2 (Delta) variant in Gauteng Province and was responsible for 98.4% of new cases sequenced in South Africa in December 2021.^8^ This fourth wave of Covid-19 arose in the context of the rollout of Covid-19 vaccines, which began on May 17, 2021. We previously conducted a population-wide seroepidemiological survey in Gauteng that was completed on January 22, 2021.^9^ We found that 19.1% of the population were anti-RBD IgG seropositive, a value that ranged from 5% to 43% across provincial sub-districts.^9^ Since that time, South Africa has experienced a third wave of Covid-19 from April 7 to November 1 that was largely due to the Delta variant, which out-competed the Beta variant.^10^

Herein, we report a follow-up seroepidemiological survey in Gauteng Province that was completed on December 9, 2021, and thus provides seroprevalence data largely from before the fourth wave. Furthermore, we report epidemiological trends for rates of Covid-19 cases, hospitalizations, recorded deaths, and excess mortality for Gauteng Province from the start of the pandemic through to January 12, 2022.

## METHODS

### Study setting and data collection

Gauteng Province is demarcated into five health districts comprising 26 sub-districts.^11^ It constitutes 1.5% of South Africa’s landmass but contains 26% (15.9/59.6 million) of its population.^11^ The overall population density (people per square kilometer) in Gauteng Province is 737, ranging from 3400 in the Johannesburg district, where 36.9% of the population live, to 200 in West Rand, in which 6.2% of the population live (Table S1 in the Supplementary Appendix).

This survey included the same households sampled during our previous survey, which was undertaken from November 4, 2020, to January 22, 2021,^9^ nine weeks after the onset of the second wave of Covid-19 in Gauteng Province, which was dominated by the Beta variant. Details of the previous survey, including the sampling framework used, have been published^9^ and are summarized in the Supplementary Methods section of the Supplementary Appendix. In the survey reported here, which was conducted from October 22 to December 9, 2021, an additional 10% of households were sampled in the same clusters to accommodate for possible non-participation, out-migration, and death of individuals since the previous survey. The survey was powered to evaluate seropositivity to SARS-CoV2 at the district and sub-district level. Demographic and epidemiologic data were collected using an electronic questionnaire,^9^ as detailed in the Supplementary Methods.

### Serology analysis

Dried blood spot samples were collected from participating individuals and tested for anti-spike and anti-nucleocapsid protein IgG (see Supplementary Methods section of the Supplementary Appendix). Anti-nucleocapsid IgG was included to identify individuals who were seropositive from natural infection rather than due to vaccination (i.e. only anti-spike IgG seropositivity). Details of the serology assay have been published and are summarized in the Supplementary Methods section of the Supplementary Appendix.^12,13^

### Covid-19 data sources

Daily case, hospital admission, and reported death data were sourced from the South African National Institute for Communicable Disease DATCOV database (latest report from January 12, 2022; see Supplementary Methods section of the Supplementary Appendix).^14^ Weekly excess death data were defined per and sourced from the South African Medical Research Council (through to January 8, 2022; see Supplementary Methods section).^15^ We analyzed these epidemiological data for Gauteng Province and its five health districts, overall and, stratified by age group and sex (where granular data available).

### Statistical analyses

Sample size justification and random household repeat sampling methods of households in our previous survey have been published^9^ and are summarized in the Supplementary Methods section of the Supplementary Appendix, together with methodology for analyses of associations with seropositivity using generalized linear models with log link to estimate risk ratios (RR). These were unadjusted, univariable analyses for each risk factor. Daily case, hospitalization, and reported death data and weekly excess death data were converted to rates using population denominators from the Statistics South Africa mid-2020 projections for South Africa and its provinces.^11^ Additional statistical methodology is summarized in the Supplementary Methods section.

### Survey Ethics

The Human Research Ethics Committee at the University of the Witwatersrand granted a waiver for ethics approval of the survey, which was being done at the behest of Gauteng Department of Health as part of public health surveillance. Neverthless, informed consent was obtained from all indviduals; those who were approached to participate were free to decline participation.

Authors designed the study, gathered and analyzed the data, vouch for the data, the analysis, and adherence to the protocol, and wrote the paper. No one who is not an author contributed to writing the manuscript.

## RESULTS

### Participants

We obtained adequate samples for serostatus evaluation from 7010 of 7498 individuals, in 3047 households (Figure 1); 83% of samples had been obtained by November 25, when the Omicron variant was first reported (Figure S1 in the Supplementary Appendix). Demographic and household characteristics, prevalence of known underlying medical conditions and self-reported HIV status, and vaccination rates are shown in Table 1. The representativeness of the survey population to the general population of Gauteng Province and of South Africa is described in Table S2 in the Supplementary Appendix. Vaccination rates in Gauteng Province by district, age, and vaccine are summarized in Table S3 in the Supplementary Appendix. As of November 25, 2021, of the total population of 12,191,569 people aged more than 12 years eligible for vaccination, 4,386,646 (36.0%) had received at least one dose of BNT162b2 or Ad26.CoV2.S, and 2,452,017 (20.1%) had received two doses. Of those aged more than 50 years, 1,074,303/2,416,045 (44.5%) had received two doses of BNT162b2.

**Table 1:** Seroprevalence of SARS-CoV-2 anti-spike or anti-nucleocapsid immunoglobulin G and risk factors for seropositivity in Gauteng Province, stratified by sex, age group, and district.

| Category | No. (%) | Seroprevalence, no. (%) [95% CI] | Risk ratio* (95%CI) |
| --- | --- | --- | --- |
| **All participants†** | 7010 (100) | 5123 (73.1) [72.0–74.1] | Not applicable |
| **Sex** | n = 7010 |  |  |
| Male | 2941 (42.0) | 1998 (67.9) [66.2–69.6] | Reference |
| Female | 4065 (58.0) | 3125 (76.9) [75.5–78.1] | 1.13 (1.10–1.17) |
| Not reported | 4 |  |  |
| **Age group – yr‡** | n = 7010 |  |  |
| <12 | 753 (10.7) | 423 (56.2) [52.6–59.7] | Reference |
| 12–18 | 622 (8.9) | 459 (73.8) [70.2–77.1] | 1.31 (1.21–1.42) |
| >18 to 50 | 4047 (57.7) | 2977 (73.6) [72.2–74.9] | 1.30 (1.23–1.40) |
| >50 | 1588 (22.7) | 1264 (79.7) [77.6–81.5] | 1.42 (1.32–1.52) |
| **Vaccination status‡** | n = 7010 |  |  |
| Not vaccinated | 5691 (81.2) | 3895 (68.4) [67.2–69.6] | Reference |
| Vaccinated | 1319 (18.8) | 1228 (93.1) [91.6–94.3] | 1.36 (1.33–1.39) |
| **Vaccination by age group** | n = 7010 |  |  |
| <12 vaccinated | 0 | 0 | Not evaluable |
| <12 unvaccinated | 753 (10.7) | 423 (55.8) [52.2–59.4] | 0.81 (0.76–0.86) |
| 12–18 unvaccinated | 603 (8.6) | 443 (73.5) [69.8–76.8] | 1.06 (1.00–1.11) |
| 12–18 vaccinated | 19 (0.3) | 16 (84.2) [60.8–94.8] | 1.21 (1.00–1.47) |
| >18 to 50 unvaccinated | 3356 (47.9) | 2334 (69.5) [68.0–71.1] | Reference |
| >18 to 50 vaccinated | 691 (9.9) | 643 (93.1) [90.9–94.7] | 1.33 (1.30–1.38) |
| >50 unvaccinated | 979 (14.0) | 695 (71.0) [68.1–73.7] | 1.02 (0.97–1.07) |
| >50 vaccinated | 609 (8.7) | 569 (93.4) [91.2–95.1] | 1.34 (1.30–1.39) |
| **Reported previous covid-19 positive test** | n = 7010 |  |  |
| Never tested | 5956 (85.0) | 4271 (71.7) [70.6–72.8] | Reference |
| Tested positive | 195 (2.8) | 172 (88.2) [82.9–92.0] | 1.23 (1.17–1.30) |
| Tested negative | 859 (12.3) | 680 (79.3) [76.3–81.8] | 1.10 (1.06–1.15) |
| **Median household members per room (IQR) – no.§** | 1 (0.5–1.5) |  | 1.01 (1.00–1.02) |
| **Occupation** | n = 7010 |  |  |
| Unemployed | 4102 (58.5) | 3014 (73.5) [72.1–74.8] | Reference |
| **Production sector** | 381 (5.4) | 279 (73.2) [68.6–77.4] | 1.00 (0.94–1.06) |
| **Teacher, public transport, retail shop** | 661 (9.4) | 509 (77.0) [73.6–80.1] | 1.05 (1.00–1.10) |
| **Healthcare worker** | 73 (1.0) | 63 (86.3) [76.4–92.5] | 1.17 (1.07–1.29) |
| **Office work/Other** | 353 (5.0) | 277 (78.5) [73.9–82.4] | 1.06 (1.01–1.13) |
| **Student** | 1440 (20.5) | 981 (68.1) [65.7–70.5] | 0.93 (0.89–0.96) |
| **Smoking status<sup>¶</sup>** | n = 5740 |  |  |
| **Non-smoker** | 4168 (59.5) | 3234 (77.6) [76.3–78.8] | Reference |
| **Daily** | 1125 (16.1) | 748 (66.5) [63.7–69.2] | 0.86 (0.82–0.90) |
| **Once or twice a week** | 244 (3.5) | 181 (74.2) [68.3–79.3] | 0.96 (0.89–1.03) |
| **Occasionally** | 203 (2.9) | 157 (77.3) [71.1–82.6] | 1.00 (0.92–1.08) |
| **Comorbidities** | n = 7010 |  |  |
| **None** | 4731 (67.5) | 3507 (74.1) [72.9–75.4] | Reference |
| **1 or more** | 2279 (32.5) | 1616 (70.9) [69.0–72.7] | 0.96 (0.93–0.99) |
| **HIV status** | n = 7010 |  |  |
| **HIV negative** | 6 460 (92.2) | 4727 (73.2) [72.1–74.2] | Reference |
| **HIV positive** | 550 (7.8) | 396 (72.0) [68.1–75.6] | 0.98 (0.93–1.04) |
| **Dwelling type<sup>¶</sup>** | n = 7010 |  |  |
| **Formal stand-alone house** | 4700 (67.0) | 3488 (74.2) [72.9–75.4] | Reference |
| **Informal dwelling** | 1147 (16.4) | 761 (66.3) [63.6–69.0] | 0.89 (0.86–0.93) |
| **Block of flats/ high-rise buildings** | 423 (6.0) | 329 (77.8) [73.6–81.5] | 1.05 (0.99–1.11) |
| **Subsidized low-income housing** | 666 (9.5) | 494 (74.3) [70.8–77.4] | 1.00 (0.95–1.05) |
| **Other** | 74 (1.1) | 51 (68.9) [57.5–78.4] | 0.93 (0.80–1.08) |
| **District** | n = 7010 |  |  |
| **Johannesburg** | 2468 (35.2) | 1880 (76.2) [74.5–77.8] | Reference |
| **Ekurhuleni** | 1861 (26.5) | 1382 (74.3) [72.2–76.2] | 0.97 (0.94–1.01) |
| **Sedibeng** | 564 (8.0) | 397 (70.4) [66.5–74.0] | 0.92 (0.87–0.98) |
| **City of Tshwane** | 1464 (20.9) | 975 (66.7) [54.2–69.0] | 0.87 (0.84–0.91) |
| **West Rand** | 653 (9.3) | 489 (74.9) [71.4–78.1] | 0.98 (0.94–1.03) |

CI, confidence interval. IQR, interquartile range. \*We determined relative risk of SARS-CoV-2 seropositivity by generalized linear models with log link to estimate risk ratios. These were unadjusted, univariable analyses for each risk factor; unadjusted risk ratios are presented with 95% confidence intervals (CI). Confidence intervals have not been adjusted for multiplicity and should not be used for inference. †Two individuals with serology results that couldn't be linked to the main questionnaire were excluded from analyses. ‡Age and vaccination status were not included in the regression model; instead, we introduced an interaction term between age and vaccination status to account for the differences in seroprevalence by vaccination status across
the different age categories. Vaccination status was obtained from vaccination certificates in 1026 of 1327 (77.3%) individuals who reported being vaccinated. <sup>§</sup>Risk ratio associated with each one additional household member per room. <sup>¶</sup>Smoking status was restricted to individuals aged >18 years. <sup>||</sup>We used the national census classification to define dwelling types.

**Figure 1:**
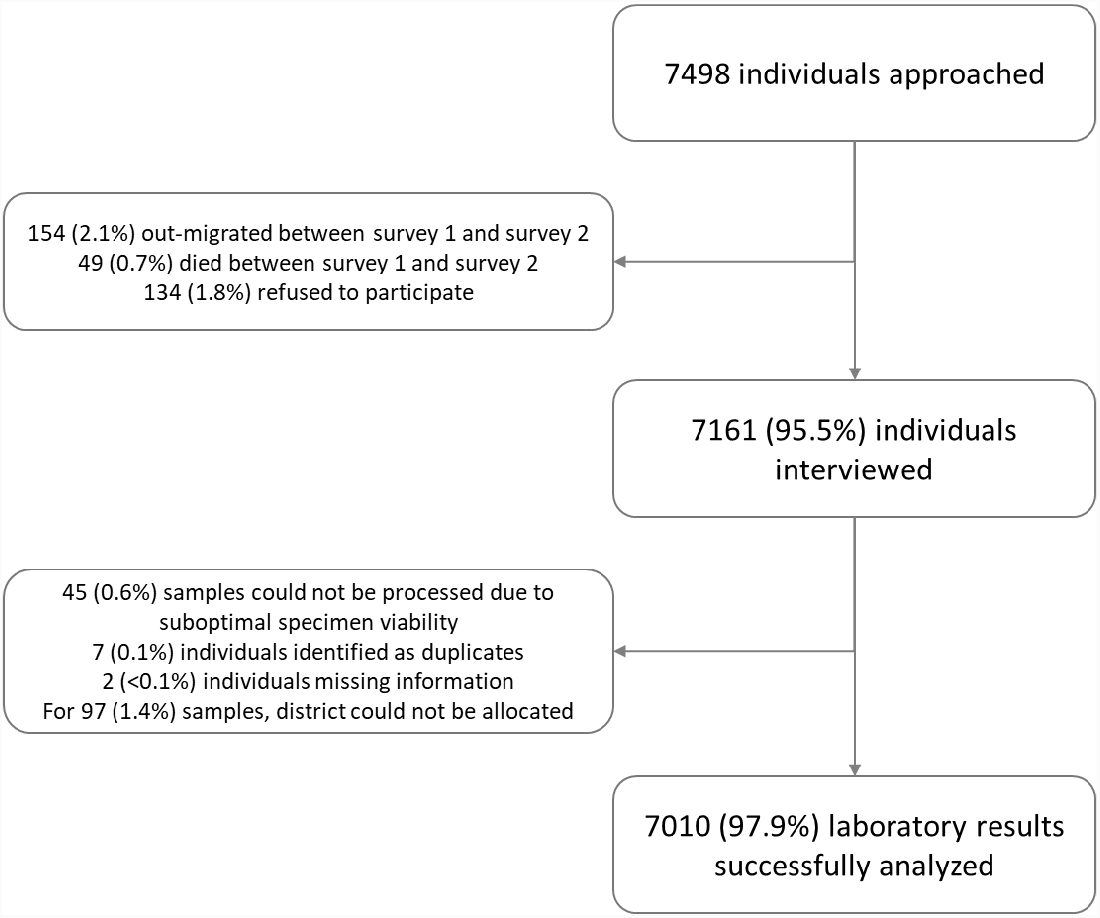
Flow of households and participants included in the seroprevalence survey. This figure illustrates the flow of participants included in the present survey (survey 2) compared to survey 1,^9^ from approaching the individuals and negotiating participation through to specimen collection and processing. Absolute numbers are presented. The final analysis included 7010 individuals in 26 sub-districts.

### Seroprevalance

In unvaccinated individuals, the overall prevalence of anti-spike or anti-nucleocapsid IgG seropositivity was 68.4% (95% confidence interval [CI], 67.2 to 69.6) (Table 1), whereas the prevalence of anti-nucleocapsid IgG seropositivity was 39.7% (2259/5691; 95% CI, 38.4 to 41.0), indicating a lack of sensitivity of anti-nucleocapsid IgG for detecting previous infection. We thus focused on the overall prevalence of anti-spike or anti-nucleocapsid IgG seropositivity.

The overall seroprevalence rate was 73.1% (95% CI, 72.0 to 74.1). Seroprevalence was heterogeneous across provincial districts (Figure S2 in the Supplementary Appendix), ranging from 66.7% (95% CI, 54.2 to 69.0) in Tshwane, where the Omicron variant was first identified, to 76.2% (95% CI, 74.5 to 77.8) in Johannesburg (Table 1). Compared with Johannesburg, seroprevalence was lower in Sedibeng and Tshwane (Table 1).

Seroprevalence was also heterogeneous at the sub-district level, with seropositivity rates ranging from 72.7% to 85.8% in Johannesburg and from 58.9% to 77.4% in City of Tshwane district (Table S4 in the Supplementary Appendix).

Females were more likely to be seropositive (76.9%) than males (67.9%; RR 1.13; 95% CI, 1.10 to 1.17) (Table 1). Seropositivity varied by age-group, being lowest in children aged less than 12 years (56.2%) and highest in individuals aged more than 50 years (79.7%). Children aged 12–18 years were more likely to be seropositive (73.8%) than those aged less than 12 years (RR 1.31; 95% CI, 1.21 to 1.42). Covid-19-vaccinated individuals were more likely to be seropositive (93.1%) than unvaccinated people (68.4%; RR 1.36; 95% CI, 1.33 to 1.39), with consistently high seropositivity across age groups and higher seropositivity in vaccinated compared with unvaccinated individuals aged 18–50 years (Table 1).

Individuals who had previously tested positive for SARS-CoV-2 infection were more likely to be seropositive (88.2%) than those who had never been tested (71.7%; RR 1.23; 95% CI, 1.17 to 1.30). Compared with participants living in a stand-alone dwelling (74.2%), participants resident in an informal settlement had a lower prevalence of seropositivity (66.3%; RR 0.89; 95% CI, 0.86 to 0.93). Daily smoking (66.5%) was associated with a lower prevalence of seropositivity compared with not smoking (77.6%; RR 0.86; 95% CI, 0.82 to 0.90) (Table 1).

### Covid-19 rates

Daily case and hospitalization, and weekly excess death rates per 100,000 population, and daily recorded death rates per 1,000,000 population in Gauteng Province are shown for the overall population in Figure 2. Daily case, hospitalization, and death rates are shown stratified by age group in Figure 3 and by sex in Figure S3 in the Supplementary Appendix. In the Omicron-dominant wave, the daily case rate increased at a faster rate and also appeared to be decreasing more quickly than in prior waves (Figure 2). Time from onset to peak of the Omicron-dominant wave was 1 month, compared with 2 months in the third wave. Although the Omicron-dominant case wave has not yet fully subsided to baseline rates before the onset of the wave, it is at its tail-end, based on the trajectory shown in Figure 2. Both recorded and excess death rates are near zero per 100,000.

**Figure 2:**
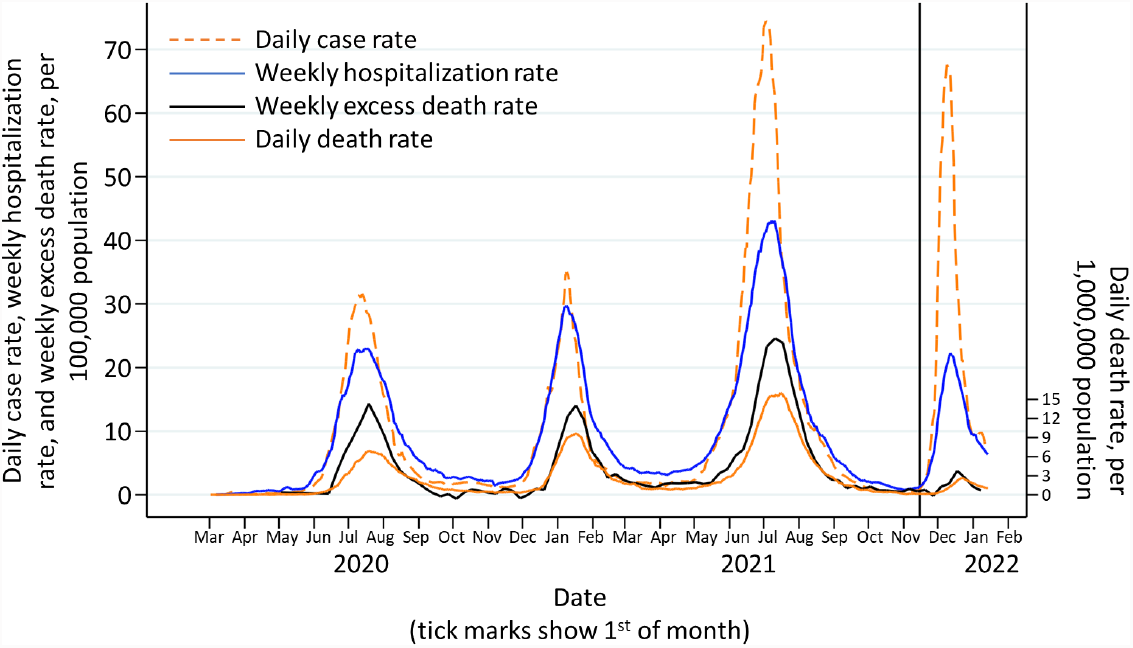
Covid-19 daily case rates, weekly hospital admission rates, weekly excess death rates, and daily reported death rates over the time period of the pandemic in Gauteng Province, South Africa, as of January 12, 2022. All data are from the National Institute for Communicable Diseases daily databases except for weekly excess deaths. Excess mortality from natural causes was defined per and sourced from the South African Medical Research Council; the excess mortality data are reported through to January 8, 2021.^15^. The solid vertical black line represents the start of the fourth, Omicron-dominant wave on November 15, 2022. Changes in testing rates, particularly the lower rates during Wave 1 due to constraints in laboratory capacity and prioritization of testing for hospitalized individuals, prevent direct comparisons, especially in terms of case numbers during the first wave in relation to the subsequent waves. Cases include asymptomatic and symptomatic individuals. Cumulative reported cases were sourced from the National Department of Health.^36^ Hospitalization data are from DATCOV, hosted by the National Institute for Communicable Disease,^14^ as described previously.^37^ The system was developed during the course of the first wave, with gradual onboarding of facilities; hence, these data could underestimate hospitalized cases in the first wave relative to subsequent waves. The hospitalized cases include individuals with Covid-19, as well as coincidental infections identified as part of routine testing for SARS-CoV-2 of individuals admitted to the facilities to assist in triaging of patients in the hospital. Cumulative reported deaths were sourced from the National Department of Health.^36^

**Figure 3:**
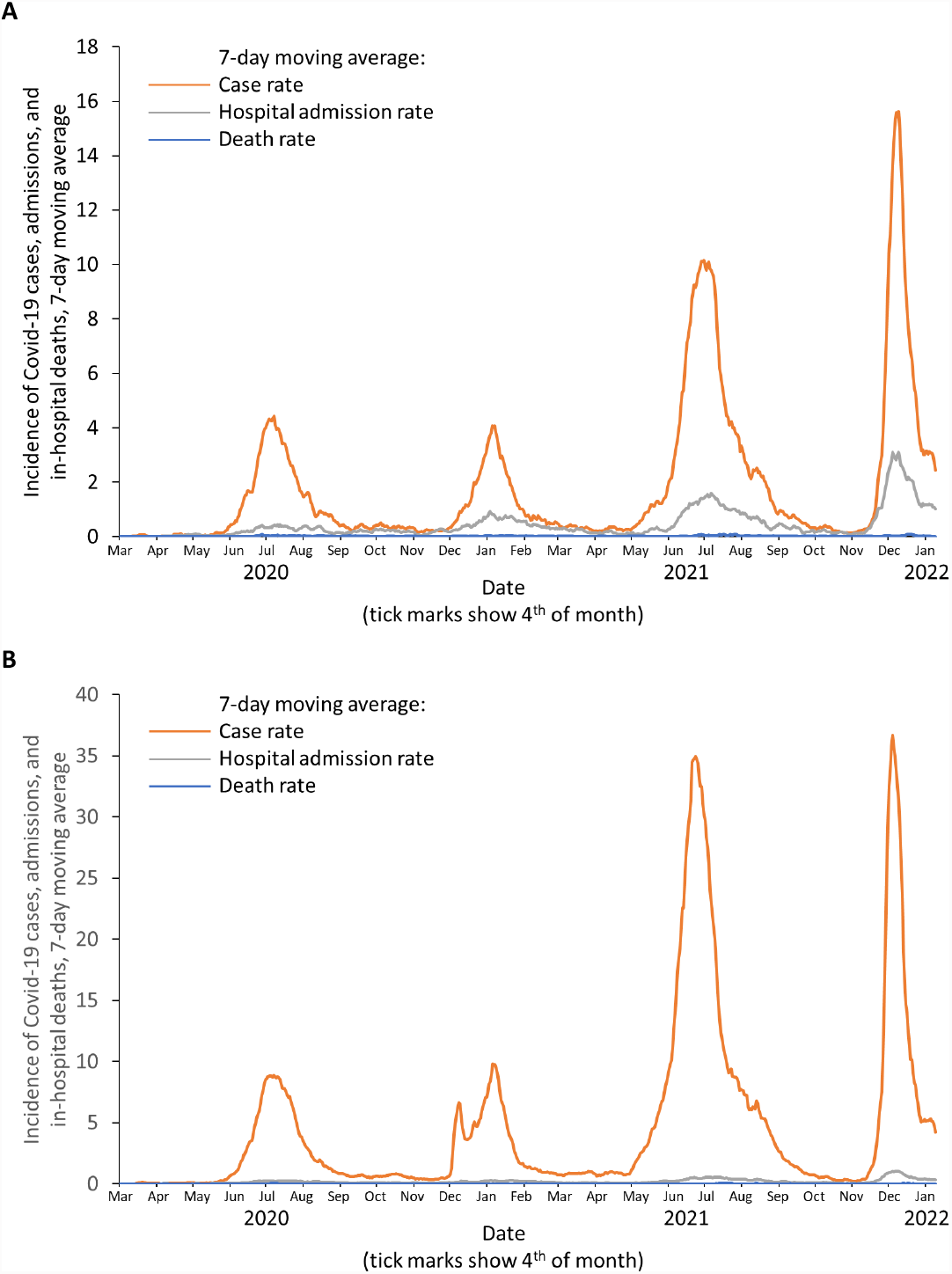
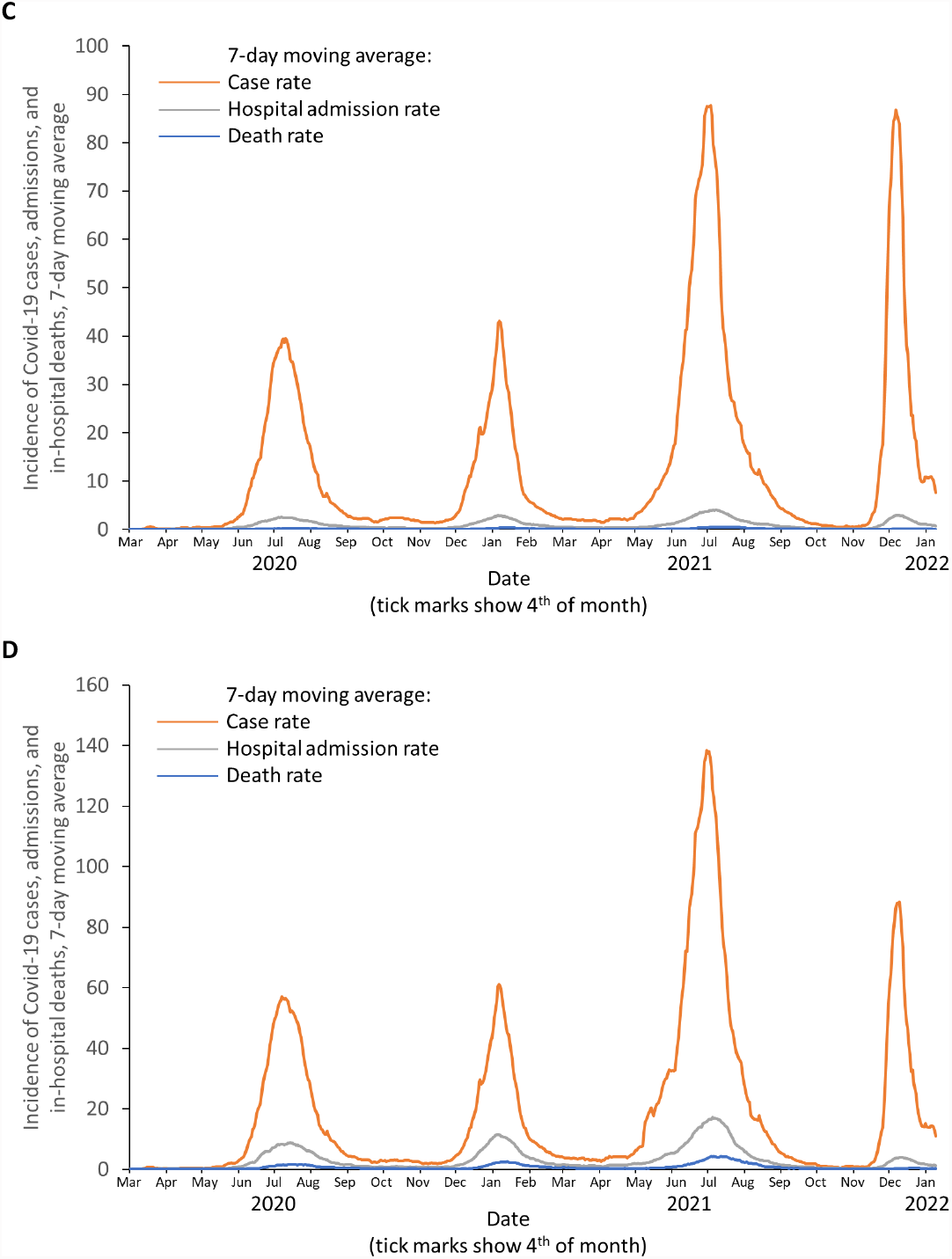
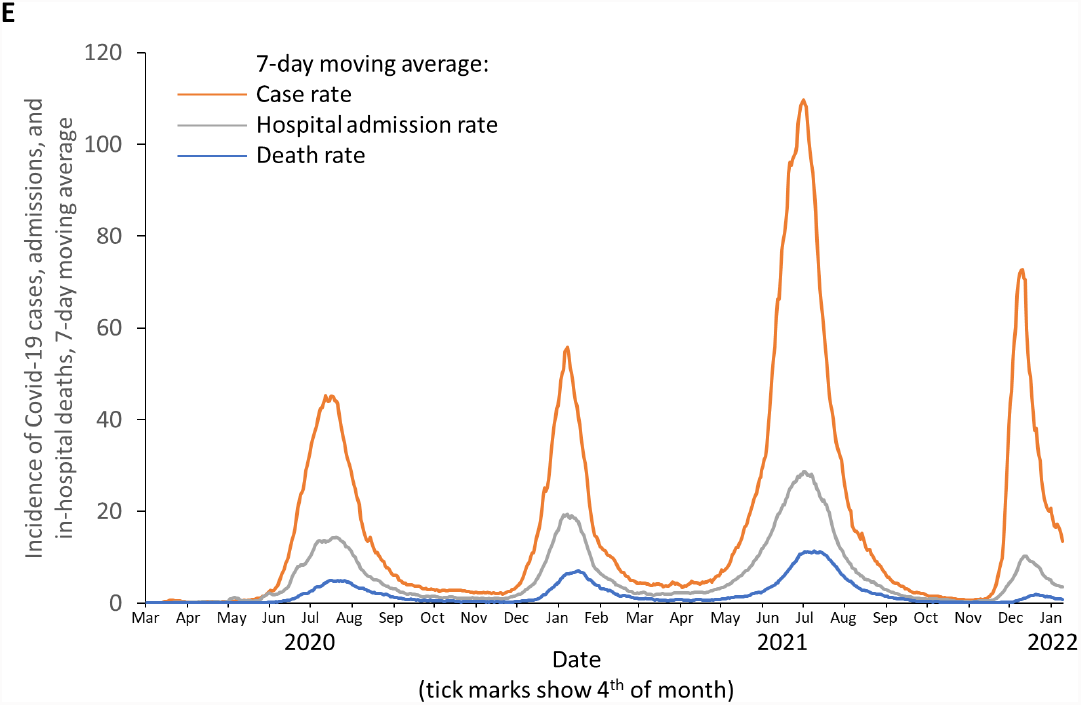
Incidence of Covid-19 cases, hospital admissions, and in-hospital deaths over the time period of the pandemic in Gauteng Province, South Africa, stratified by age group. Panels show data for individuals aged (A) 0–4 years, (B) 5–17 years, (C) 18–44 years, (D) 45–59 years, and (E) 60 years and older. As absolute rates differ between age groups, different Y-axis scales have been used for each individual age group in order to provide clarity and aid visual interpretation of the trends in each age group.

The number of documented Covid-19 cases in the Omicron-dominant case wave (n=226,932) was higher compared with wave 2 (n=182,564) and lower than in wave 3 (n=511,638), whereas rates of hospitalizations, recorded deaths, and Covid-19 attributable excess mortality were consistently lower than in earlier waves (Table 2). The peak incidence rates for hospitalization, recorded deaths, and excess mortality in the fourth wave were also lower than in previous waves (Figure 2, Table 2). The Omicron-dominant wave contributed 11.2%, 3.9%, and 3.3% of overall Covid-19 hospitalizations, recorded deaths, and excess mortality, respectively, compared with 43.6%, 49.3%, and 52.7%, respectively, in the Delta-dominant third wave (Table 2). Similar trends were observed across all districts (Figure S4). Although a lag in excess death reporting exists (January 8, 2022), the current rate of 12 per 100,000 is lower than 197 per 100,000 recorded in the third wave (Figure 2). Rates are on an ongoing downward trajectory, with 7-day-moving-average incidence rates (per 100,000) of 7.3 cases, 0.96 hospitalizations, and 0.11 deaths on January 12, 2022 (representing 9.3-fold, 3.3-fold, and 2.4-fold reductions compared to the peak rates of 67.6, 3.18, and 0.26, respectively), and are nearing pre-wave baseline rates (respectively, 0.46, 0.15, and 0.04 on October 25, 2021).

**Table 2:** Cumulative reported Covid-19 cases, hospitalizations, recorded deaths, and excess mortality in Gauteng Province by Covid-19 wave.

|  | **Wave 1\*** | **Wave 2\*** | **Wave 3\*** | **Wave 4\*** | **TOTAL** |
| --- | --- | --- | --- | --- | --- |
| **Dominant variant** | Wild type | Beta | Delta | Omicron |  |
| **Cases** |  |  |  |  |  |
| **Period of wave** | Mar 7–Nov 13, 2020 | Nov 14, 2020–Mar 30, 2021 | Mar 31–Oct 25, 2021 | Oct 26, 2021–Jan 12, 2022 | Mar 7, 2020–Jan 12, 2022 |
| **Cases in wave – no.†** | 232,130 | 182,564 | 511,638 | 226,932 | 1,153,264 |
| **Case rate per 100,000 population** | 1498 | 1178 | 3301 | 1464 | 7440 |
| **Proportion of total cumulative cases, %** | 20.1 | 15.8 | 44.4 | 19.7 | 100 |
| **Hospitalizations** |  |  |  |  |  |
| **Period of wave** | Mar 7–Nov 6, 2020 | Nov 7, 2020–Apr 6, 2021 | Apr 7–Oct 31, 2021 | Nov 1, 2021–Jan 12, 2022 | Mar 7, 2020–Jan 12, 2022 |
| **Hospitalizations in wave – no.‡** | 33,315 | 30,685 | 61,642 | 15,789 | 141,431 |
| **Hospitalization rate per 100,000 population** | 215 | 198 | 398 | 102 | 912 |
| **Proportion of total cumulative hospitalizations, %** | 23.6 | 21.7 | 43.6 | 11.2 | 100 |
| **Reported deaths** |  |  |  |  |  |
| **Period of wave** | Mar 31–Dec 14, 2020 | Dec 15, 2020–May 2, 2021 | May 3–Nov 19, 2021 | Nov 20, 2021–Jan 12, 2022 | Mar 31, 2020–Jan 12, 2022 |
| **Reported deaths in wave – no.** | 6443 | 7084 | 14,256 | 1116 | 28,899 |
| **Reported death rate per 100,000 population§** | 42 | 46 | 92 | 7 | 186 |
| **Proportion of total cumulative reported deaths, %** | 22.3 | 24.5 | 49.3 | 3.9 | 100 |
| **Excess deaths** |  |  |  |  |  |
| **Period of wave** | May 9–Dec 19, 2020 | Dec 20, 2020–Mar 26, 2021 | Mar 27–Nov 25, 2021 | Nov 26, 2021–Jan 8, 2022 | May 9, 2020–Jan 8, 2022 |
| **Excess deaths in wave – no.** | 13,476 | 11,970 | 30,546 | 1,927 | 57,919 |
| **Excess death rate per 100,000 population** | 87 | 77 | 197 | 12 | 374 |
| **Proportion of total cumulative excess deaths, %** | 23.3 | 20.7 | 52.7 | 3.3 | 100 |

\*All data are from the National Institute for Communicable Diseases daily databases except for weekly excess deaths. Excess mortality from natural causes was defined per and sourced from the South African Medical Research Council; the excess mortality data are reported through to January 1, 2021.<sup>15</sup> Other waves are lagged with respect to cases. Consequently, each of the hospitalization, recorded death, and excess death waves have their own cut-points determining the start and end of the 4 waves. The Omicron-dominant fourth case wave is at its tail-end but has not yet fully subsided. Totals, incidence, and proportions of cases, hospitalizations, deaths, and excess deaths are anticipated to continue to increase somewhat over the next few weeks until the respective waves have fully subsided.
†Changes in testing rates, particularly the lower rates during Wave 1 due to constraints in laboratory capacity and prioritization of testing for hospitalized individuals, prevent direct comparisons, especially in terms of case numbers during the first wave in relation to the subsequent waves. Cases include asymptomatic and symptomatic individuals. Cumulative reported cases were sourced from the National Department of Health.<sup>36</sup>
\*Hospitalization data are from DATCOV, hosted by the National Institute for Communicable Disease,<sup>14</sup> as described previously.<sup>37</sup> The system was developed during the course of the first wave, with gradual onboarding of facilities; hence, these data could underestimate hospitalized cases in the first wave relative to subsequent waves. The hospitalized cases include individuals with Covid-19, as well as coincidental infections identified as part of routine testing for SARS-CoV-2 of individuals admitted to the facilities to assist in triaging of patients in the hospital.
<sup>§</sup>Cumulative reported deaths were sourced from the National Department of Health.<sup>36</sup>

The lower incidence of hospitalizations and recorded deaths during the Omicron-dominant wave was evident across all age groups older than 19 years and when stratified by sex. In contrast, the incidence of hospitalizations and recorded deaths during the fourth wave in children aged less than 19 years, which were generally markedly lower than in older age groups, were relatively unchanged compared with earlier waves, except for a lower death rate in the group aged 5–19 years compared with during the Delta-dominant third wave (Figure 3, Supplementary Tables S5, S6, and S7).

## DISCUSSION

The resurgence of Covid-19 in Gauteng Province dominated by the Omicron variant evolved at a time when Covid-19 vaccine coverage was 36.0% in people aged more than 12 years, with only 20.1% having received at least two doses of a Covid-19 vaccine as part of the national vaccine roll-out program. Nevertheless, our survey shows widespread underlying SARS-CoV-2 seropositivity across the province (73.1%), including up to 85.8% in some sub-districts, before the onset of the current Omicron-dominant wave. This high rate of seropositivity has been primarily induced by prior SARS-CoV-2 infection, as evidenced by the 68.4% seropositivity rate in Covid-19-unvaccinated individuals. The random sampling methodology used for selecting households in the serosurvey, which was proportionated to the sub-district population size, ensures representativeness to the general population of Gauteng Province.

In this context, we observed a dramatic decoupling of hospitalization and death rates from infection rate compared with previous waves. The biological basis for this decoupling is possibly the extensive cell-mediated immunity in the population induced by previous natural infection and vaccination, for which coverage of at least one dose was 61.2% (1,479,288/2,416,045) in adults aged more than 50 years (who accounted for 81% [22,269/27,500] of all deaths in Gauteng Province through to the end of the third wave^16^). Although we did not evaluate cell-mediated immunity, other studies have reported that natural infection induces a diverse polyepitopic cell-mediated immune response targeted against the spike protein, nucleocapsid protein, and membrane protein.^17^ Consequently, cell-mediated immunity is likely more durable than neutralizing antibody-mediated immunity in the context of small mutations,^18^ particularly those mainly affecting the spike protein, as in the Omicron variant. Furthermore, natural infection induces robust memory T-cell responses, including long-lived cytotoxic (CD8^+^) T-cells, which have a half-life of 125–255 days.^19^ We believe that the evolution of cell-mediated immunity from prior natural infection and vaccination has resulted in the decoupling of the high case rates seen with the Omicron variant and the rates of severe disease. This is despite the Omicron variant evading neutralizing antibody activity induced by spike-protein-based vaccines and by prior infections with other variants not harboring the same full set of putatively antibody-evasive mutations. Our hypothesis is supported by two recent preprint publications indicating that the majority of the T-cell response induced by vaccination or natural infection cross-recognizes the Omicron variant, thereby likely contributing to protection against severe disease.^20,21^ An alternative or additional mechanism by which protection against severe disease may be conferred, despite reduced neutralizing antibody activity against the Omicron variant, is through Fc-mediated effector functions of non-neutralizing antibodies inducing antibody-mediated cellular phagocytosis, complement deposition, and natural killer cell activation.^18,22^ In addition, the Omicron variant may be less potent at causing serious illness.

We saw high Covid-19 case rates due to the Omicron variant despite the high seropositivity prevalence of humoral immune responses, consistent with the Omicron variant being antibody-evasive. Reports indicate that the Omicron variant is more evasive to neutralizing antibody activity than even the Beta variant.^7,23-25^ Relative to vaccine-induced neutralizing antibody activity against WT virus, neutralizing activity after two doses of BNT162b2 or AZD1222 (ChAdOx1 nCoV-19) is reduced substantially.^26,27^ Nevertheless, the majority of individuals with hybrid immunity from natural infection and BNT162b2 or AZD1222 vaccination have measurable neutralizing activity against the Omicron variant, albeit lower than against the WT virus.^23^ In this context, the high rate of breakthrough cases and reinfections with the Omicron variant is to be expected in South Africa, where the majority of individuals have developed immunity from natural infection, which induces lower-magnitude anti-spike neutralizing and binding antibody responses compared with vaccination.^24^ Furthermore, South Africa only provided a single dose of Ad26.COV2.S as part of its vaccine rollout at the time of the evolution of the fourth wave, which induces lower neutralizing and blocking antibody titers than two doses of the BNT162b2,^24^ and third doses of BNT162b2 had yet to be introduced in South Africa at that time.

This clinical evidence of the antibody-evasiveness of the Omicron variant is corroborated by early studies reporting limited vaccine efficacy (VE) against Omicron at 25 weeks after two doses of AZD1222 or BNT162b2.^28^ However, VE increased substantially at 2 weeks after a booster dose of BNT162b2,^28^ which results in much higher neutralizing antibody titers than after two doses of vaccine^29^ and thus may partly mitigate the relative antibody-evasiveness of the Omicron variant. Similarly, in South Africa, vaccine effectiveness against hospitalization was 70%, compared to 93% observed against the Delta variant.^30^ These data, together with the very limited neutralizing antibody activity against the Omicron variant following two doses of AZD1222 or BNT162b2, further corroborate that protection against severe Covid-19 is likely mediated by much lower neutralizing antibody titres^24^ or primarily through cell-mediated immunity and/or non-neutralizing antibodies with Fc-effector functions.^18,22^

Analogous to the emerging experience with the Omicron variant is the antibody-evasiveness of the Beta variant in recipients of AZD1222, the Astra-Zeneca chimpanzee adenovirus-based vaccine, which showed no efficacy against mild-to-moderate Covid-19 due to the Beta variant.^31^ However, effectiveness of 80% against hospitalization or death due to the Beta or Gamma variants has been reported from Canada.^32^ While AZD1222 induced nominal neutralizing antibody activity against the Beta variant, only 11 of the 87 spike-protein epitopes targeted by T-cell immune responses induced by AZD1222 were affected by mutations in the Beta variant.^31^ The dissociation between the lack of AZD1222-induced neutralizing antibody activity and protection against severe lower respiratory tract disease was also observed in a challenge study with AZD122 against the Beta variant in a Syrian golden hamster model.^33^

The greater transmissibility of the Omicron variant is corroborated by the rapid rise of reported Covid-19 cases in Gauteng Province during the course of the fourth wave. Indeed, the rate of increase in cases exceeds any of the previous three waves, indicating that the Omicron variant is more transmissible than even the Delta variant, which has an estimated reproductive rate (Ro) of 5-6.^34^

Limitations of our study include the use of publically available data on Covid-19 morbidity and mortality that were collated in surveillance systems and could have changed over time, which could affect comparisons across the four waves. DATCOV surveillance does not distinguish between SARS-CoV-2 cases hospitalized for Covid-19 and those admitted for other illness who coincidentally test positive for SARS-CoV-2 on routine screening. Nevertheless, these systems are unlikely to have changed since the Delta-dominant third wave. Differences in overall testing rates over time also limits head-to-head comparions of case rates between the waves, albeit that criteria for testing have been similar since the start of the second wave. Another limitation is that the Omicron-dominant case wave has not fully subsided to the incidence observed before its onset. The numbers and proportions of total cumulative cases, hospitalizations, deaths, and excess deaths attributable to this wave are anticipated to continue to increase somewhat over the next few weeks until the respective waves have fully subsided, in particular the data for hospitalizations and deaths due to the time lag following infections. Also, our contention that cell-mediated immunity primarily due to natural infection, with or without Covid-19 vaccination, is resulting in the decoupling of case rates and severe disease remains to be investigated. In particular, the extent to which the polyepitopic T-cell response induced by vaccination against the spike-protein and the even more diverse polyepitopic T-cell response stimulated by natural infection, with or without vaccination, remain cross-reactive against the Omicron variant warrants further investigation.^20,21^

Another possible contributing factor to the decoupling of cases and severe disease rates with the Omicron variant compared with previous variants is that the Omicron variant may be more adept at infecting the upper airways and less adept at infecting the lower airways, which could result in reduced virulence.^35^ The differing prevalence of immunity at the time of Omicron-dominant wave compared with in previous waves limits our ability to draw any conclusions on the relative roles of reduced virulence and higher prevalence of underlying cell-mediated immunity in contributing to the decoupling of infection and severe Covid-19 rates observed with the Omicron variant in our study.

We believe that the decoupling of case rates compared with hospitalization and death rates experienced with the Omicron-dominant wave in South Africa heralds a turning point in the Covid-19 pandemic, if the primary goal is protection against severe disease and death rather than trying to prevent infections. The 70% effectiveness seen with BNT162b2 against severe disease in South Africa^30^ might well be due to the hybrid cell-mediated immunity induced by vaccination and natural infection. Whether the same protection against severe Covid-19 due to the Omicron variant will be seen in countries in which immunity is mainly from vaccination remains to be determined.

## Data Availability

Data are available at www.wits-vida.org; requests for data sharing should be directed to Professor Shabir A. Madhi

## ACKNOWLEDGMENTS

Funding support for the seroepidemiological survey was provided by the Bill & Melinda Gates Foundation (grant number: INV-023514). DATCOV, as a national surveillance system, is funded by the National Institute for Communicable Diseases (NICD) and the South African National Government.

The authors thank: Andronica Moipone Shonhiwa, Genevie Ntshoe, Joy Ebonwu, Lactatia Motsuku, Liliwe Shuping, Mazvita Muchengeti, Jackie Kleynhans, Gillian Hunt, Victor Odhiambo Olago, Husna Ismail, Nevashan Govender, Ann Mathews, Vivien Essel, Veerle Msimang, Tendesayi Kufa-Chakezha, Nkengafac Villyen Motaze, Natalie Mayet, Tebogo Mmaborwa Matjokotja, Mzimasi Neti, Tracy Arendse, Teresa Lamola, Itumeleng Matiea, Darren Muganhiri, Babongile Ndlovu, Khuliso Ravhuhali, Emelda Ramutshila, Salaminah Mhlanga, Akhona Mzoneli, Nimesh Naran, Trisha Whitbread, Mpho Moeti, Chidozie Iwu, Eva Mathatha, Fhatuwani Gavhi, Masingita Makamu, Matimba Makhubele, Simbulele Mdleleni, Bracha Chiger, Jackie Kleynhans, Michelle Groome from the the Epidemiology team; and Tsumbedzo Mukange, Trevor Bell, Lincoln Darwin, Fazil McKenna, Ndivhuwo Munava, Muzammil Raza Bano, and Themba Ngobeni from the Information Technology team, at the National Institute for Communicable Diseases (NICD) Notifiable Medical Conditions Surveillance System (NMCSS).

The authors would like to acknowledge Steve Hill, PhD, of Ashfield MedComms, an Ashfield Health company, part of UDG Healthcare, for medical writing and editing support that was funded by AstraZeneca in accordance with Good Publication Practice (GPP3) guidelines (Ann Intern Med. 2015;163:461–4). Gauteng Department of Health is acknowledged for providing the Vaccine Coverage Data.

---

**Figure 2:**
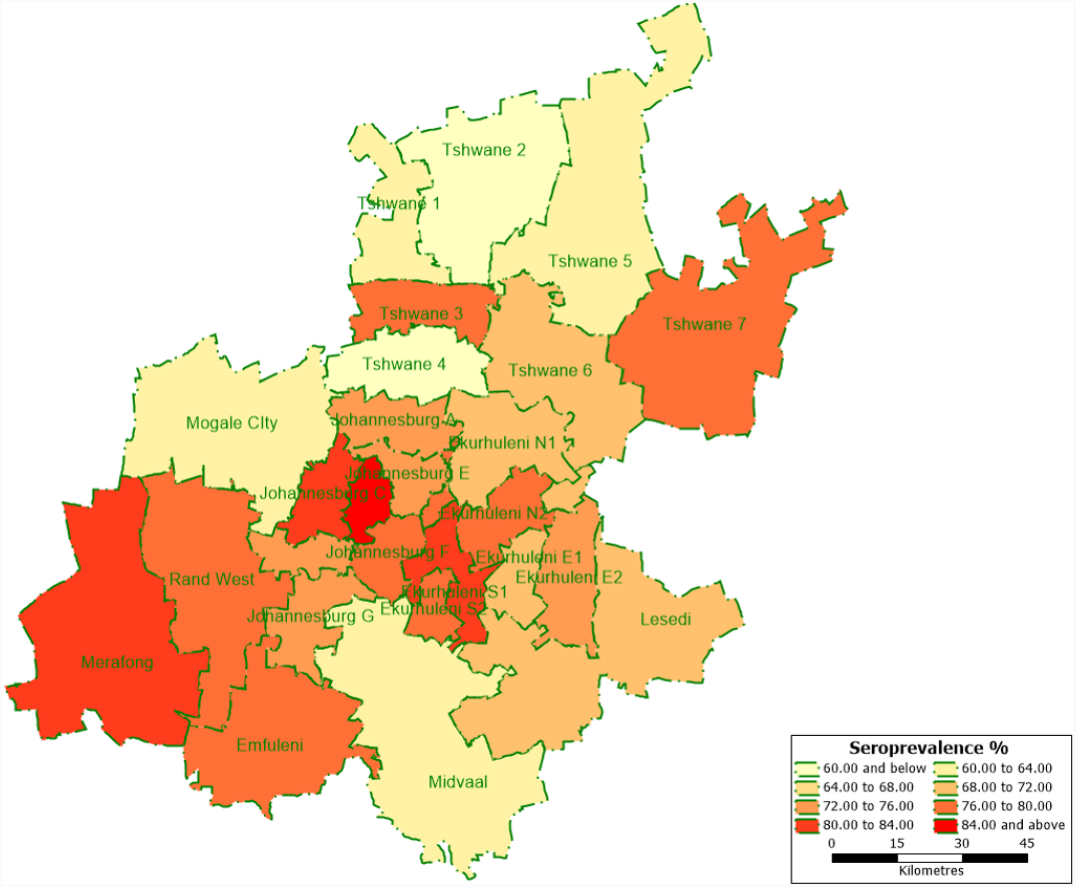
Seroprevalence across sub-districts in Gauteng Province. Sampling period from October 22, 2021 through to December 9, 2021.

**Figure 3:**
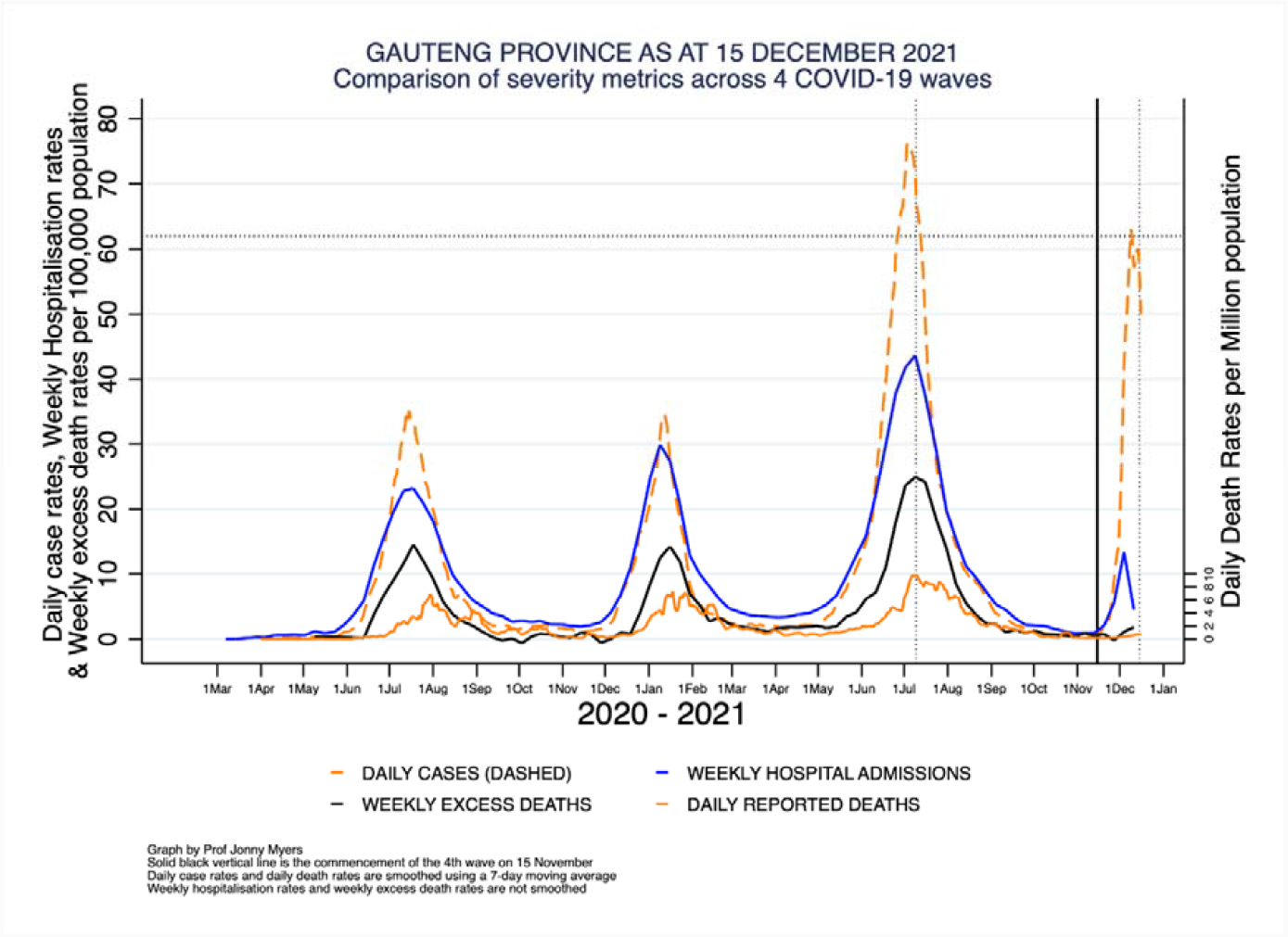
Covid-19 daily case rates, weekly hospital admission rates, weekly excess death rates, and daily reported death rates over the time period of the pandemic in Gauteng Province, South Africa. The daily rates are smoothed using a 7-day moving average while the weekly rates are unsmoothed. Hospital admissions data in Gauteng Province were provided by the National Institute of Communicable Diseases.^15^Daily case and daily death data were sourced from the South African national coronavirus database produced by the National Department of Health.^13^Weekly excess death data were sourced from the South African Medical Research Council weekly report for epidemiological week 48.^14^

