## Supplemental material for "South African Population Immunity and Severe Covid-19 with Omicron Variant"

**Supplementary Appendix**


| **Contents** | **Page** |
|---|---|
| **Supplementary Methods** | **3** |
| Study design and sample size | 3 |
| Data collection | 3 |
| Sample collection and processing | 3 |
| Serology analysis | 4 |
| Covid-19 data sources | 5 |
| Statistical analyses | 6 |
| **Supplementary Figures** | **8** |
| Figure S1: Cumulative number and percentage of dried blood spots collected from November 22, 2021 to December 9, 2021 | 8 |
| Figure S2: Seroprevalence across sub-districts in Gauteng Province. | 9 |
| Figure S3: Incidence of Covid-19 cases, hospital admissions, and in-hospital deaths over the time period of the pandemic in Gauteng Province, South Africa, stratified by sex | 10 |
| Figure S4: Covid-19 daily case rates, weekly hospital admission rates, weekly excess death rates, and daily reported death rates over the time period of the pandemic in the five districts of Gauteng Province, South Africa, as of January 12, 2022 | 11 |
| **Supplementary Tables** | **14** |
| Table S1: Demographics of sampling area in serosurvey | 14 |
| Table S2. Representativeness of the seroepidemiological survey population to the general population of Gauteng Province and of South Africa | 15 |
| Table S3: Vaccination rates in Gauteng Province, South Africa, by district, age group, and vaccine received, as of November 25, 2021 | 16 |
| Table S4: Seroprevalence of SARS-CoV-2 anti-spike (anti-S) or anti-nucleocapsid (anti-N) immunoglobulin G (IgG) Gauteng Province across the Districts and sub-Districts | 17 |
| Table S5. Number and incidence per 100,000 population of Covid-19 cases, in Gauteng Province by Covid-19 wave, stratified by age and by gender | 19 |
| Table S6. Number and incidence per 100,000 population of hospitalizations in Gauteng Province by Covid-19 wave, stratified by age and by gender | 20 |
| Table S7. Number and incidence per 100,000 population of recorded deaths in Gauteng Province by Covid-19 wave, stratified by age and by gender | 21 |
| **Supplementary References** | **22** |

### Supplementary Methods

#### Study design and sample size

This survey included households that had been sampled during the first seroprevalence survey, which was conducted done from November 4, 2020, to January 22, 2021,[1] and included an additional 10% households per cluster to accommodate for non-participation. The sample size calculation was based on the Africa Centres for Disease Control and Prevention generic protocol for a population-based, age- and gender-stratified seroprevalence survey study for SARS-CoV-2[2] and on the World Health Organization population-based age-stratified seroepidemiological investigation protocol for COVID-19 virus infection.[3] Assuming a seroprevalence of 10%, response rate of 0.75, intra-cluster correlation (ICC) of 0.33, with a precision of 0.1, α of 0.05, and design effect of 3.31, the resultant required overall survey sample size was 6025 (61 to 1948 by sub-district) individuals. Design effect of 3.31 was based on the observation that seroprevalence rates vary by geographic area within the same region.[2] A conservative ICC of 0.33 was used considering clustering nature of respiratory diseases.

Three-stage sampling was employed; first, the GeoTeraImage (GTI)-2019 dataset (https://geoterraimage.com/), which comprises more than 16,000 "small areas" used for demarcating census areas was stratified by housing type. Systematic random cluster selection without replacement and probability proportional to estimated size (PPES) was then used to select clusters. Finally, a random sample of 9 households was selected from each cluster. The number of clusters, and hence the number of households selected per sub-district, was proportional to the sub-district population size. The average household size was assumed to be 4 individuals based on census data with a target of 9 households per cluster. All individuals residing in sampled households, irrespective of age, were eligible.

#### Data collection

Electronic data collection was done using android tablets with real time synchronisation of data into a central KoBo Collect database. The questionnaire comprised a household module with household membership listing and socio-demographic characteristics. The individual module included individual-level questions on socio-demographics, previous Covid-19 diagnosis, Covid-19 vaccination, comorbidities, and health seeking behaviour (see Supplementary Appendix 2).

#### Sample collection and processing

Dried blood spots (DBS) were obtained by finger-prick with a single-use lancet needle, with 3–5 DBS collected on filter cards (Ahistrom Munksjo, Germany, catalogue number 8.460.0013.A). The DBS were dried for 3 hours at room temperature, packed into plastic pouches with silica-gel sachets, transported to the laboratory, and stored at –20°C until analysis.[1] Elution of antibodies from DBS card specimens was performed as previously described.[4] Briefly, one spot was cut from the filter card using a 6 mm hole punch and added to 600 µl of assay buffer. The spot was kept in a shaker at 2–8°C overnight for elution and the following day was centrifuged at 2000 g for 10 min before analysis.

**Serology analysis**

SARS-CoV-2 full-length spike and nucleocapsid (N) protein immunoglobulin G (IgG) were measured by quantitative assay on the Luminex platform. The expression plasmid encoding for the SARS-CoV-2 full-length spike were obtained from Florian Krammer, Mount Sinai, USA. The recombinant spike protein was expressed as described previously,[5] and N protein was obtained from BioTech, Africa (Cat no# BA25-P, South Africa). Proteins were coupled to magnetic microsphere beads (Bio-Rad, USA) using a two-step carbodiimide reaction.[6] Samples were analyzed in single-plex, and each plate included two in-house control sera. Bead fluorescence was read with the Bio-Plex 200 instrument (Bio-Rad) using Bio-Plex manager 5.0 software (Bio-Rad).

An in-house reference serum was developed by pooling convalescent serum from adult SARS-CoV-2-positive patients. This interim reference serum was calibrated against research reagent NIBSC 20/130 and the first WHO International Standard for anti-SARS-CoV-2 NIBSC 20/136. Binding antibody unit (BAU) values assigned to the in-house reference serum were 2819 BAU/mL for spike IgG and 1101 BAU/mL for N protein IgG. The assay was further evaluated for detection of antibodies against SARS-CoV-2 using Covid-19 convalescent plasma panel NIBSC 20/118.

The IgG assay using DBS cards was validated using paired plasma and DBS cards (collected from hospitalized participants who were SARS-CoV-2-positive by PCR test) for detection of SARS-CoV-2 IgG antibodies. There was a good correlation between IgG values measured by the two sampling methods: $r = 0.935$ and $r = 0.965$ for receptor binding domain and full-length spike protein of SARS-CoV-2, respectively. Bland-Altman assessment showed good agreement between two sampling methods.[4]

Serum samples collected prior to 2020 ($n = 31$) and serum samples obtained from randomly selected ($n = 15$) participants who were SARS-CoV-2-positive on PCR test and who had serial blood sampling before and after symptom onset (including individuals with mild-to-moderate illness and asymptomatic infections) were used for the analysis of assay specificity and sensitivity, and for calculation of the threshold for IgG seropositivity. Based on IgG titers from pre-Covid-19 era samples, baseline and post-infection samples of participants who were SARS-CoV-2-positive on PCR test, 32 BAU/mL and 15

BAU/mL were selected as the thresholds indicative of seropositivity for full-length spike and N protein IgG, respectively. The sensitivity of the assay to detect seropositivity at the selected thresholds was 100% (15/15) and 85% (12/14) for samples taken >14 days following the SARS-CoV-2-positive PCR test for full-length spike and N protein IgG, respectively.

**Covid-19 data sources**

Daily case, hospital admission, and reported death data were sourced from the DATCOV database, which is hosted by South Africa's National Institute for Communicable Diseases,[7,8] as described previously.[9] The system was developed during the course of the first wave of Covid-19, with gradual onboarding of facilities. Hence, data from DATCOV could underestimate hospitalized cases in the first wave relative to subsequent waves. The hospitalized cases include individuals with Covid-19 as well as coincidental infections identified as part of routine testing for SARS-CoV-2 of individuals admitted to the facilities to assist in triaging of patients in the hospital.

DATCOV was initially implemented in eight sentinel public hospital sites but within six months was adopted as South Africa's national Covid-19 hospital surveillance system. In addition, the South African National Department of Health placed data capturers at all public hospitals in 8 provinces (excluding Western Cape, which has an electronic information system that feeds into DATCOV). This has ensured the quality and completeness of data submission of DATCOV. By January 7, 2021, 481,209 admissions and 97,931 deaths had been reported to DATCOV from 259 private and 407 public hospitals in South Africa. It is believed that the reporting of Covid-19 admissions to DATCOV is near-complete and representative of Covid-19 admissions across the country.

DATCOV contains data on all individuals, irrespective of age, who had a positive reverse transcription polymerase chain reaction (RT-PCR) assay or antigen test for SARS-CoV-2, with a confirmed duration of stay in hospital of one full day or longer, regardless of reason for admission. The DATCOV case report form (CRF) was adapted from the World Health Organization (WHO) COVID-19 case reporting tool,[10] recording the following variables: demographic data (age, sex, race), exposures such as occupation, and potential risk factors such as comorbid disease(s) and pregnancy status. Additional variables included level of treatment, complications, treatment, and outcomes of the hospital admission (discharged, transferred out to another hospital, or died).

Data on excess weekly mortality were sourced from the weekly report by the Burden of Disease Research Unit of the South African Medical Research Council.[11] Within the glossary to each report, the following statements are provided regarding the definition and calculation of excess mortality rates:

- "Excess deaths: There is no universal definition of, or understanding of what is meant by, "excess mortality". It is a term used in epidemiology and public health that refers to the number of deaths that are occurring above what we would normally expect. The WHO uses the term to describe "Mortality above what would be expected based on the non-crisis mortality rate in the population of interest. Excess mortality is thus mortality that is attributable to the crisis conditions. It can be expressed as a rate (the difference between observed and non-crisis mortality rates), or as a total number of excess deaths."
- "Excess natural deaths associated with COVID-19: Generally, the number of excess deaths per week is calculated as the number of all-cause deaths in that week less the number that might be assumed to have occurred had there not been the epidemic (i.e. the counterfactual number), provided that the counterfactual is lower. However, this approach has generally only been applied to countries where deaths have been tracking the counterfactual before the onset of significant numbers of COVID-19 related deaths. The method provides a poor estimate of the numbers of COVID-19 and collateral deaths in the early stages of the epidemic when this is not the case. Thus, we estimated the numbers of COVID-19 and collateral deaths, once a clear upward trend is evident, as the number of actual deaths less a baseline number determined as a proportion of the predicted number. By the end of the 1st wave of the pandemic, the predicted values have been used as the counterfactual."

**Statistical analyses**

We estimated anti-SARS-CoV-2 seroprevalence as the proportion of individuals testing positive for either spike or N protein IgG and assessed variability by age, gender, vaccination status, and sub-district of residence. Factors associated with seropositivity were determined using generalized linear models with log link to estimate risk ratios. These were unadjusted, univariable analyses for each risk factor. Confidence intervals have not been adjusted for multiplicity and should not be used for inference. Comorbidities included any of self-reported hypertension, diabetes, asthma, HIV positive status, cancer, tuberculosis, stroke, or lung, liver, kidney, or heart disease. Stata (version 16.1) was used for data analyses, with sampling weights calculated based on the sampling frame. Data analyses techniques used clustering adjusted for at the household level.

For epidemiological analyses, daily rates were smoothed using locally weighted scatterplot smoothing (Lowess) in STATA version 16.1 using a bandwidth of 0.06, which closely matches the 7-day moving average but which provides a smoother graph line, allowing the minimum values at the beginning and end of each Covid-19 wave to be easily identified. These minima are taken as cut-points for each wave, with no overlap and no data omitted from analysis. Raw counts were obtained at these cut-points for cumulative numbers for each wave. Weekly excess death rates were unsmoothed. Data on excess mortality were unavailable by age group and sex.

**Figure S1: Cumulative number and percentage of dried blood spots collected from November 22, 2021 to December 9, 2021.**

By November 25, when the Omicron variant was first reported, 83% of dried blood spot (DBS) samples had been collected.

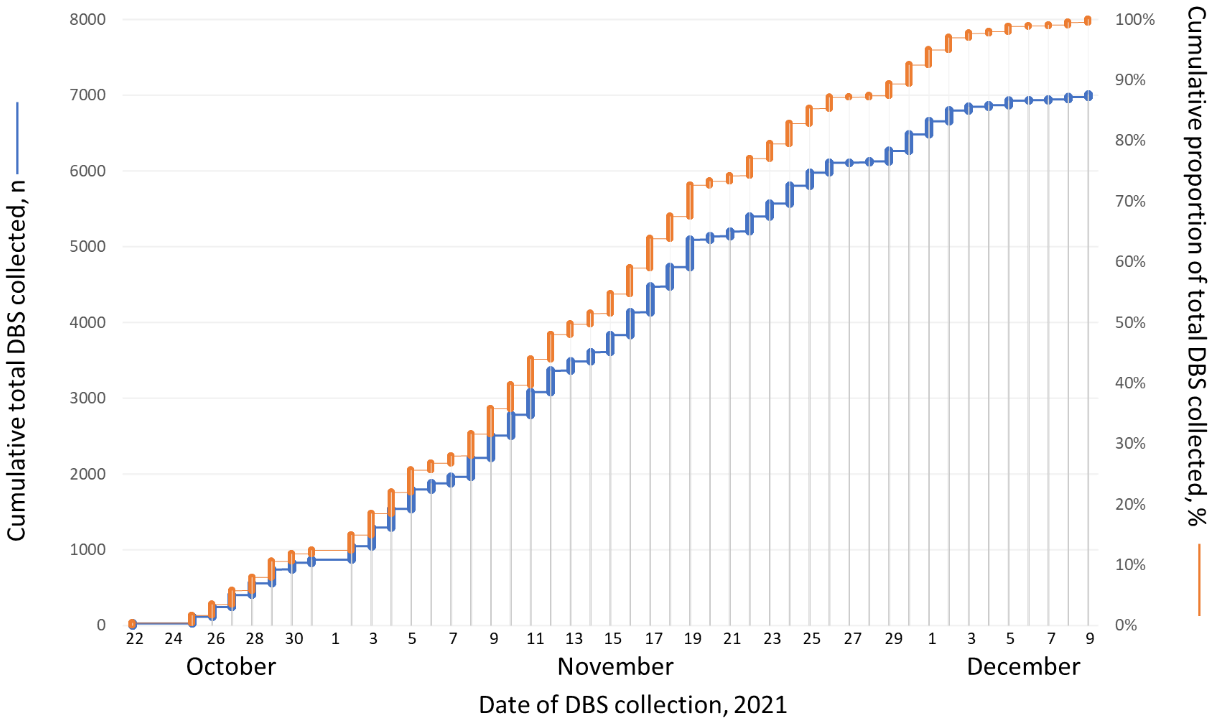

***Figure S2:* Seroprevalence across sub-districts in Gauteng Province.**
Sampling period from October 22, 2021 through to December 9, 2021.

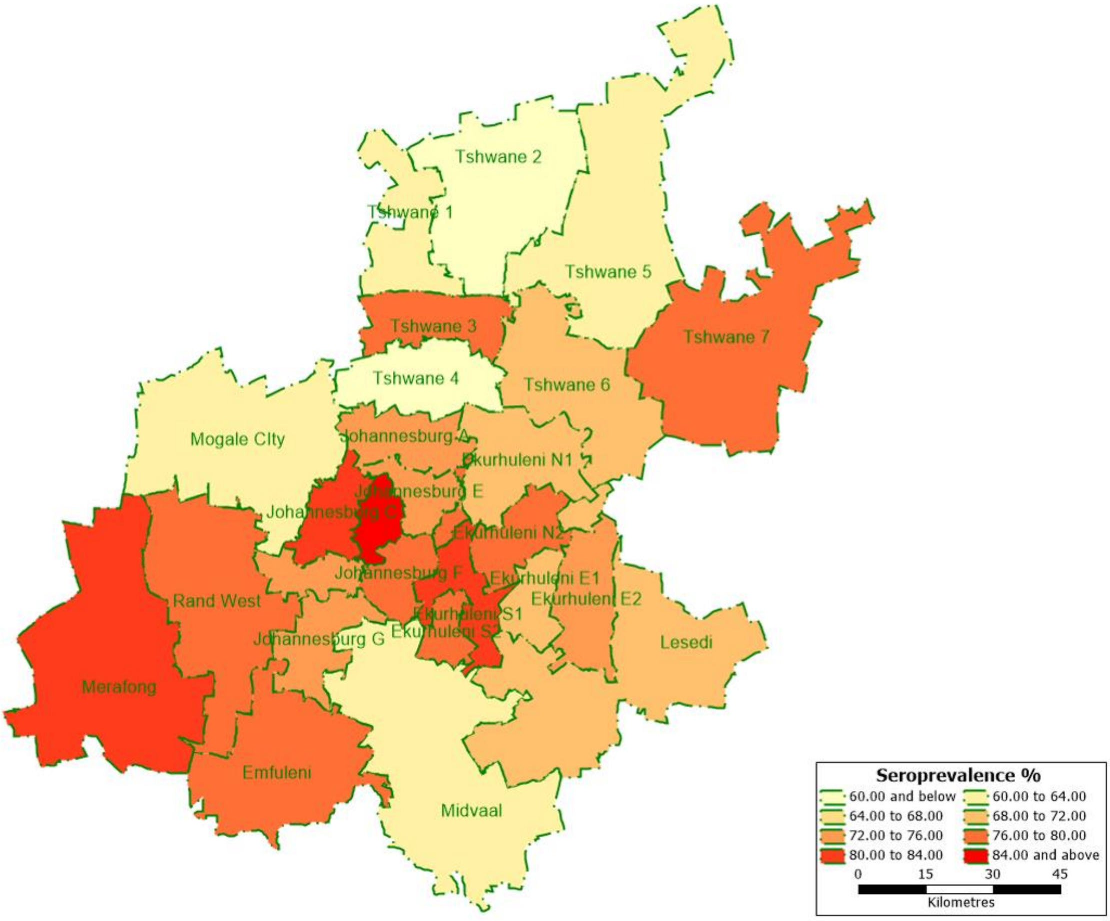

***Figure S3:*** **Incidence of Covid-19 cases, hospital admissions, and in-hospital deaths over the time period of the pandemic in Gauteng Province, South Africa, stratified by sex**
Panels show data for (A) males and (B) females.

**A**

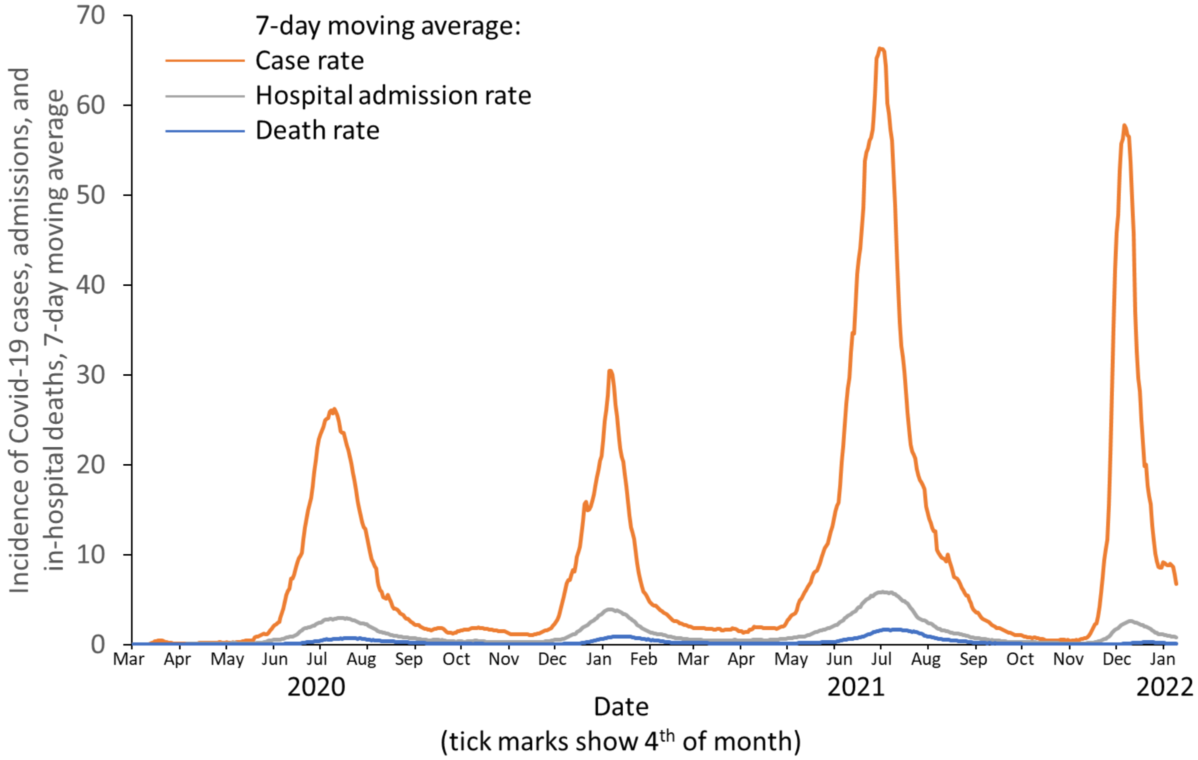


**B**

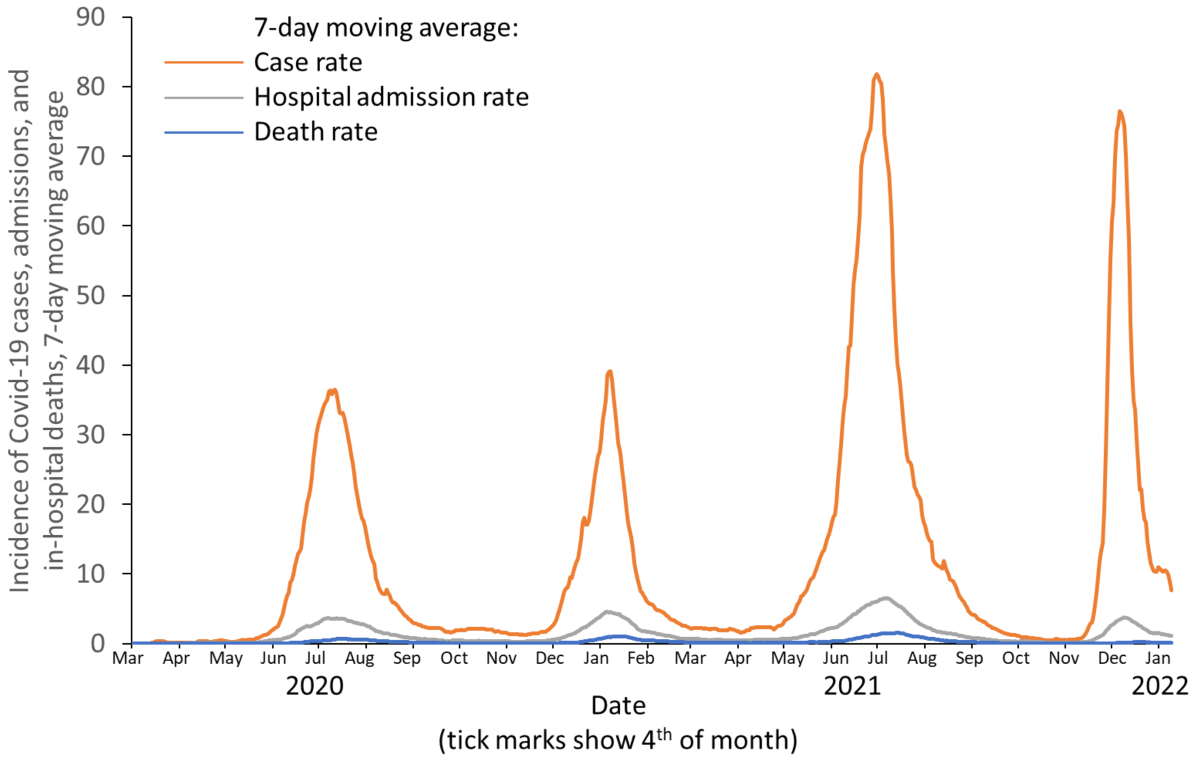

***Figure S4:* Covid-19 daily case rates, weekly hospital admission rates, weekly excess death rates, and daily reported death rates over the time period of the pandemic in the five districts of Gauteng Province, South Africa, as of January 12, 2022**

All data are from the National Institute for Communicable Diseases daily databases except for weekly excess deaths. Excess mortality from natural causes was defined per and sourced from the South African Medical Research Council; the excess mortality data are reported through to January 8, 2021.[11] Stratified excess mortality data were unavailable for Sedibeng and West Rand.[11] The solid vertical black line represents the start of the fourth, Omicron-dominant wave on November 15, 2022. Changes in testing rates, particularly the lower rates during Wave 1 due to constraints in laboratory capacity and prioritization of testing for hospitalized individuals, prevent direct comparisons, especially in terms of case numbers during the first wave in relation to the subsequent waves. Cases include asymptomatic and symptomatic individuals. Cumulative reported cases were sourced from the National Department of Health.[12] Hospitalization data are from DATCOV, hosted by the National Institute for Communicable Disease,[7] as described previously.[9] The system was developed during the course of the first wave, with gradual onboarding of facilities; hence, these data could underestimate hospitalized cases in the first wave relative to subsequent waves. The hospitalized cases include individuals with Covid-19, as well as coincidental infections identified as part of routine testing for SARS-CoV-2 of individuals admitted to the facilities to assist in triaging of patients in the hospital. Cumulative reported deaths were sourced from the National Department of Health.[12] As absolute rates differ between districts, different Y-axis scales have been used for each individual district in order to provide clarity and aid visual interpretation of the trends in each district.

**Johannesburg District**

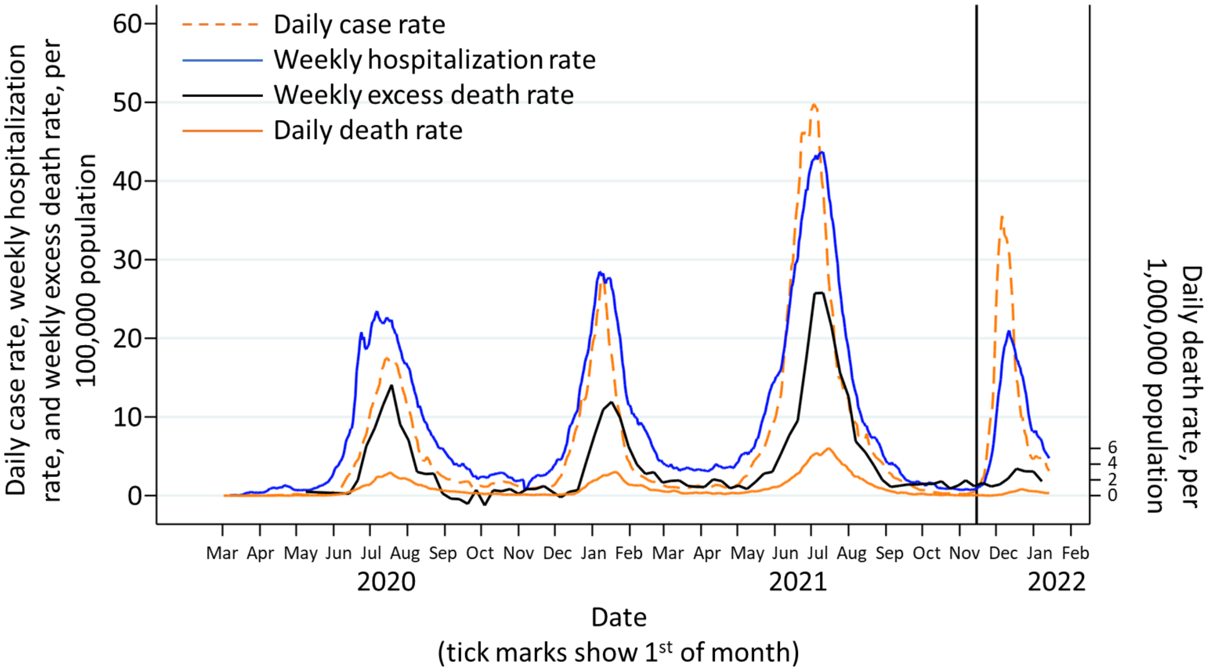

**Ekurhuleni District**

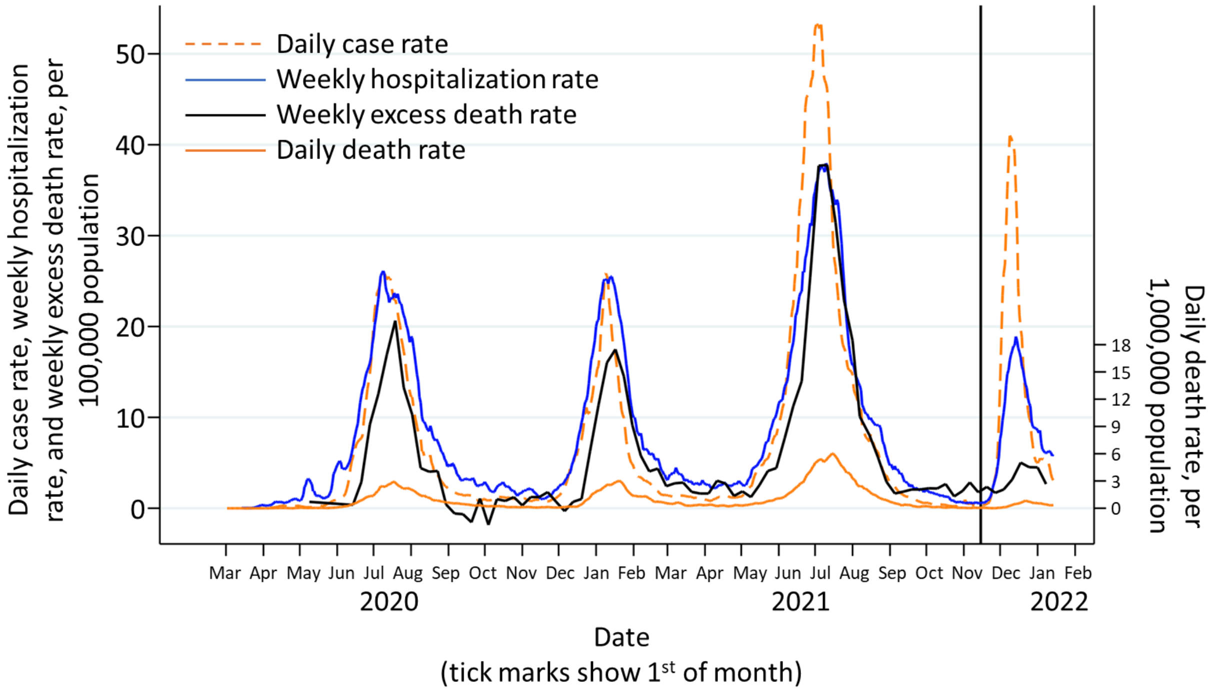


**Sedibeng District**

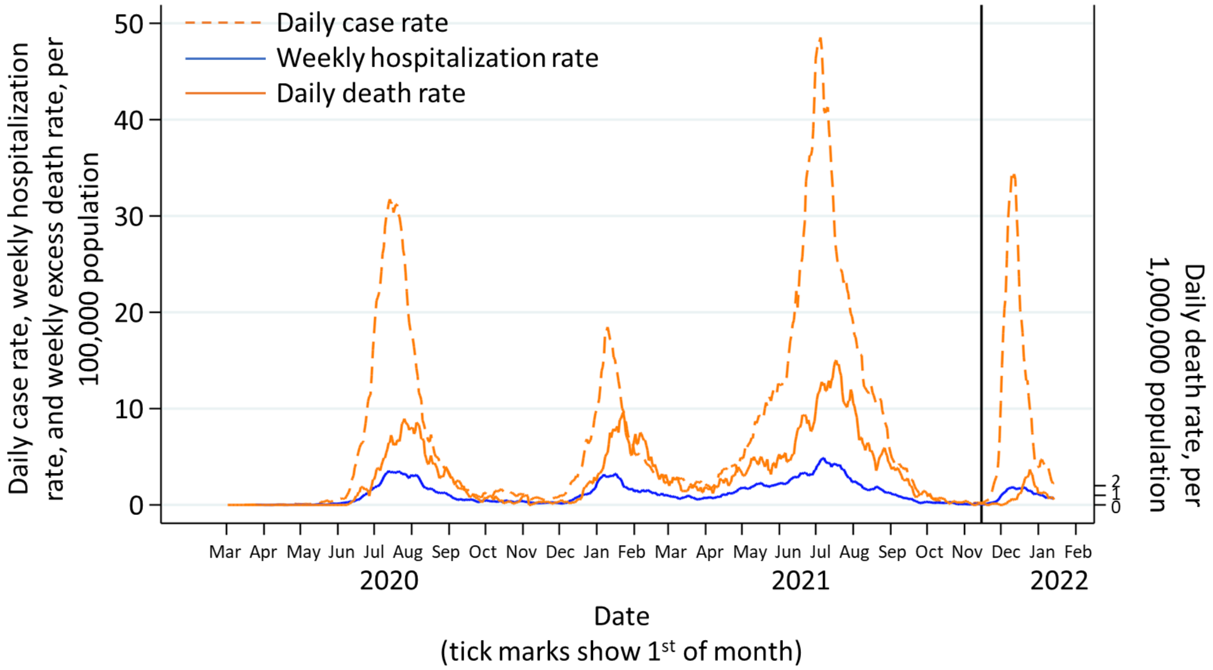

**City of Tshwane District**

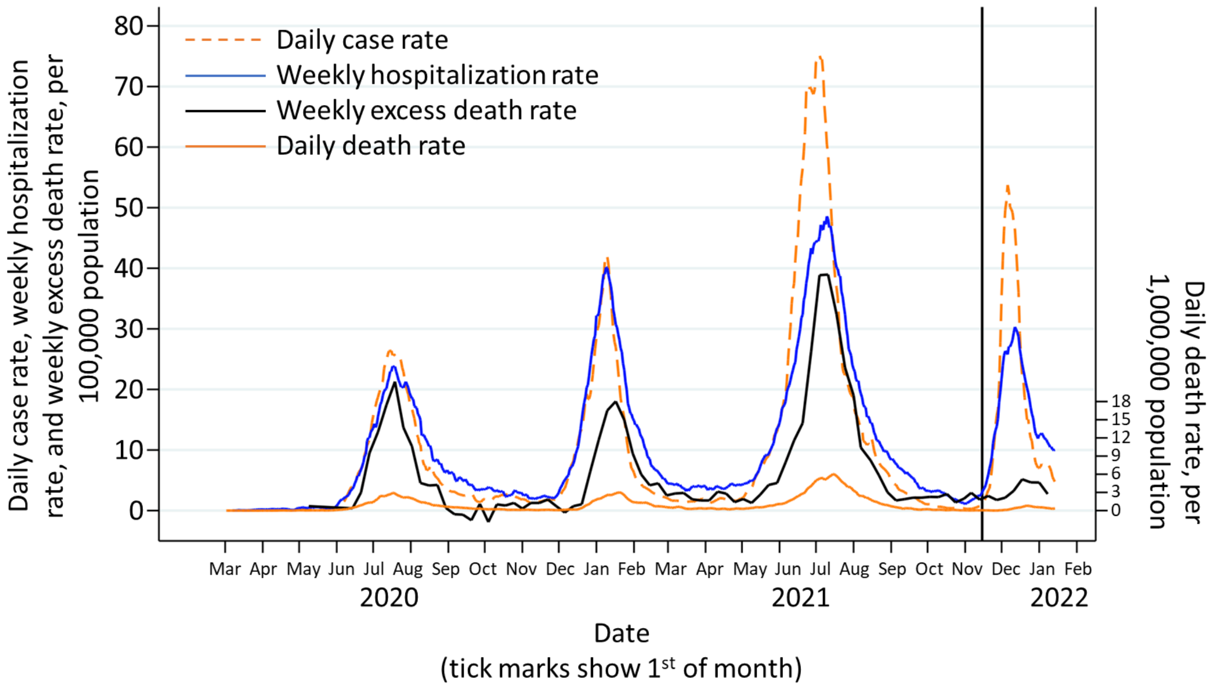


**West Rand District**

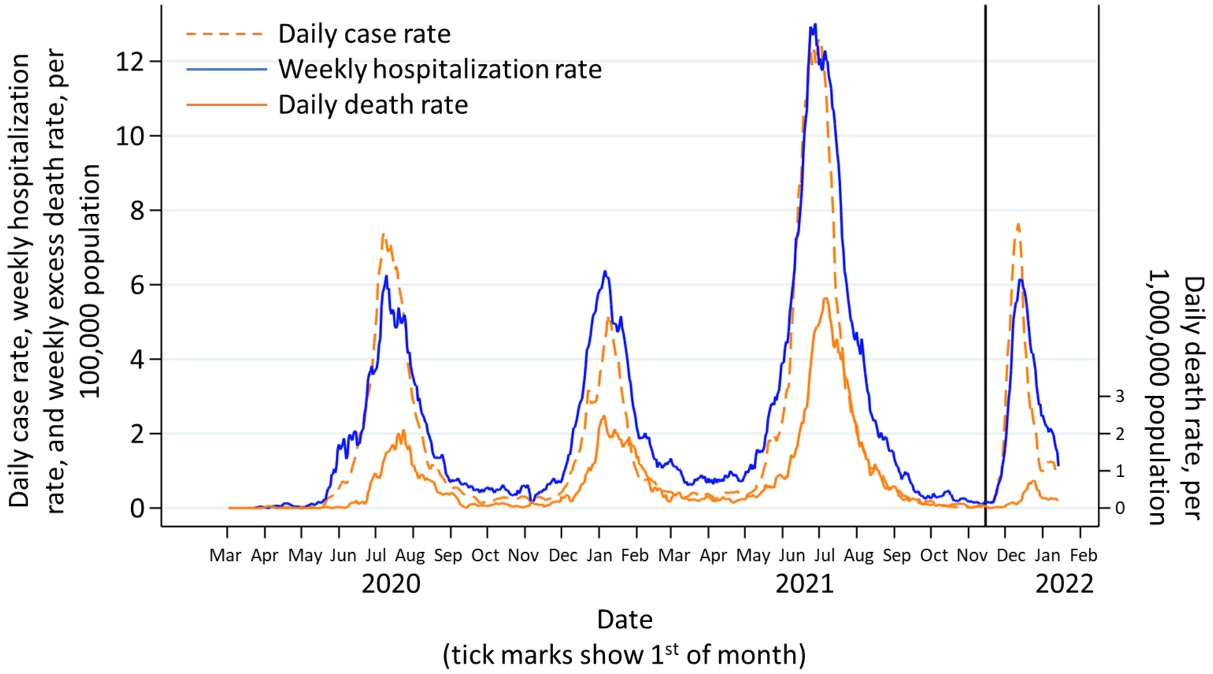

**Table S1: Demographics of sampling area in serosurvey.**

| District / Sub-district | Population – no.* | Participants in survey | Survey time-period, 2021 | Population density – persons/km$^2$† | Living in informal housing settlement – %† |
|---|---|---|---|---|---|
| **Gauteng Province** | **15,176,113** | **7010** | **Oct 22–Dec 09** | **737** | **20.1%** |
| **Johannesburg District** | **5,606,238** | **2468** | **Oct 22–Dec09** | **3400** | **18.6%** |
| Johannesburg A | 779,519 | 333 | Oct 25–Nov 23 | | |
| Johannesburg B | 435,241 | 197 | Nov 16–25 | | |
| Johannesburg C | 799,980 | 444 | Oct 25–Dec 08 | | |
| Johannesburg D | 1,396,243 | 646 | Oct 22–Dec 09 | | |
| Johannesburg E | 601,433 | 161 | Nov 23–Dec 08 | | |
| Johannesburg F | 751,484 | 243 | Oct 27–Dec 03 | | |
| Johannesburg G | 842,339 | 444 | Oct25–Dec03 | | |
| **Ekurhuleni District** | **3,825,650** | **1861** | **Oct 22–Dec 08** | **1609** | **22.6%** |
| Ekurhuleni E1 | 626,517 | 353 | Oct 25–Dec 06 | | |
| Ekurhuleni E2 | 455,262 | 252 | Oct 22–Dec 08 | | |
| Ekurhuleni N1 | 708,290 | 358 | Nov 05–30 | | |
| Ekurhuleni N2 | 697,175 | 258 | Nov 16–Dec 05 | | |
| Ekurhuleni S1 | 673,758 | 210 | Nov 07–Dec 02 | | |
| Ekurhuleni S2 | 664,648 | 430 | Oct 25–Dec 08 | | |
| **Sedibeng District** | **1,084,503** | **564** | **Oct 25–Dec 03** | **220** | **16.3%** |
| Emfuleni | 127,419 | 408 | Oct 25–Dec 03 | | |
| Lesedi | 126,285 | 104 | Nov 16–23 | | |
| Midvaal | 830,798 | 52 | Nov 09–26 | | |
| **City of Tshwane District** | **3,709,635** | **1464** | **Oct 25–Dec 09** | **464** | **19.3%** |
| Tshwane 1 | 1,032,885 | 471 | Oct 28–Dec 09 | | |
| Tshwane 2 | 436,950 | 175 | Nov 04–25 | | |
| Tshwane 3 | 730,788 | 229 | Nov 03–Dec 05 | | |
| Tshwane 4 | 482,448 | 78 | Oct 26–Dec 01 | | |
| Tshwane 5 | 119,190 | 204 | Oct 25–Dec 06 | | |
| Tshwane 6 | 768,446 | 245 | Nov 12–Dec 07 | | |
| Tshwane 7 | 138,928 | 62 | Nov 29–Dec 08 | | |
| **West Rand District** | **950,088** | **653** | **Oct 25–Dec 09** | **200** | **23.9%** |
| Mogale City | 435,254 | 149 | Oct 25–Dec 09 | | |
| Rand West City | 300,960 | 261 | Nov 03–Dec 09 | | |
| Merafong City | 213,874 | 243 | Nov 02–Dec 09 | | |

*Population estimates obtained from the STATS-SA provincial mid-year population estimates.
†Only available at the district level.

**Table S2.** Representativeness of the seroepidemiological survey population to the general population of Gauteng Province and of South Africa[13]

| **Disease** | **Coronavirus disease 2019 (Covid-19)** |
|---|---|
| Special considerations related to: | |
| Sex and gender | Covid-19 case rates are slightly higher in females compared to males |
| Age | Covid-19 incidence (cases per 100,000 persons) increases with age; the lowest incidence is in those aged <5years (862.4), and incidence increases linearly with increase in age, with the highest incidence in those aged 50–54 years (11,160.6). Covid-19 incidence in those aged >60 years is lower than in those aged 20–59 years |
| Race or ethnic group | There is no evidence that Covid-19 disproportionately affects people of a specific race or ethnicity. In South Africa, 81% of the population is Black African. |
| Geography | Covid-19 incidence varies by geographic area.[1,13,14] Gauteng Province has the highest incidence rate in South Africa, accounting for 33% of the total cases in the country. Across the world, Covid-19 incidence and severity vary widely associated with underlying population structures; countries with older populations have more Covid-19 cases. |
| Socio-economic status | In two seroprevalence surveys conducted in Gauteng Province, inclusive of this one, there was lower seroprevalence in informal settlements compared to formal stand-alone houses and higher seroprevalence in blocks of flats/high-rise buildings.[1] In Gauteng Province, 20.1% of the population resides in informal settlements,[15] ranging from 16.3% to 23.9% across the five districts. Similarly, of our survey sample, 16.4% reside in informal settlements. |
| Overall representativeness of this survey | In South Africa and Gauteng Province, females constitute 51.2% and 49.9% of the population, respectively. Biological gender at birth was reported by the participants and captured as male, female, or declined to respond. This survey comprised 58% females, and thus slightly overrepresented females. Covid-19 incidence is higher in older populations and Africa generally has younger populations compared to Europe and America. 73.6% of this survey population were aged 15–59 years comparable to 67.7% in Gauteng Province and South Africa. Our survey sample was representative of the underlying population residential types, an indicator for socio-economic status. Seroprevalence estimates by age and gender from this survey are consistent with Covid-19 epidemiological data in South Africa.[13] |

**Table S3: Vaccination rates in Gauteng Province, South Africa, by district, age group, and vaccine received, as of November 25, 2021.**

| Place | Total target population – no. | At least one vaccine dose – no. (%) | Two doses of either vaccine – no. (%) | One dose of BNT162b2* – no. (%) | Two doses of BNT162b2* – no. (%) | One dose of Ad26.COV2.S† – no. (%) | Sisonke2 study Ad26.COV2.S booster† – no. (%) |
|---|---|---|---|---|---|---|---|
| **Gauteng Province** | | | | | | | |
| **Overall** | **12,191,569** | **4,386,646 (36.0)** | **2,452,017 (20.1)** | **3,261,822 (26.8)** | **2,417,414 (19.8)** | **1,124,824 (9.2)** | **34,603 (0.3)** |
| 12–18 yr | 1,447,321 | 158,646 (11.0) | 35,993 (2.5) | 139,691 (9.7) | 35,933 (2.5) | 18,955 (1.3) | 60 (<0.1) |
| >18 to 50 yr | 8,328,203 | 2,748,011 (33.0) | 1,329,064 (16.0) | 1,880,401 (22.6) | 1,306,882 (15.7) | 867,610 (10.4) | 22,182 (0.3) |
| >50 yr | 2,416,045 | 1,479,288 (61.2) | 1,086,664 (45.0) | 1,241,139 (51.4) | 1,074,303 (44.5) | 238,149 (9.9) | 12,361 (0.5) |
| Not defined | n/a | 701 | 296 | 591 | 296 | 110 | 0 |
| **Johannesburg** | | | | | | | |
| **Overall** | **4,643,131** | **1,879,851 (40.5)** | **1,097,108 (23.6)** | **1,470,587 (31.7)** | **1,074,606 (23.1)** | **409,264 (8.8)** | **22,502 (0.5)** |
| 12–18 yr | 525,964 | 71,743 (13.6) | 17,180 (3.3) | 65,326 (12.4) | 17,137 (3.3) | 6417 (1.2) | 43 (<0.1) |
| >18 to 50 yr | 3,251,303 | 1,220,750 (37.5) | 651,704 (20.0) | 905,004 (27.8) | 637,022 (19.6) | 315,746 (9.7) | 14,682 (0.5) |
| >50 yr | 865,864 | 586,946 (67.8) | 428,063 (49.4) | 499,909 (57.7) | 420,286 (48.5) | 87,037 (10.1) | 7,777 (0.9) |
| Not defined | n/a | 412 | 161 | 348 | 161 | 64 | 0 |
| **Ekurhuleni** | | | | | | | |
| **Overall** | **3,090,769** | **960,276 (31.1)** | **555,245 (18.0)** | **749,337 (24.2)** | **551,445 (17.8)** | **210,939 (6.8)** | **3800 (0.1)** |
| 12–18 yr | 368,349 | 35,077 (9.5) | 8264 (2.2) | 32,255 (8.8) | 8261 (2.2) | 2822 (0.8) | 3 (<0.1) |
| >18 to 50 yr | 2,115,789 | 595,824 (28.2) | 289,862 (13.7) | 423,366 (20.0) | 287,462 (13.6) | 172,458 (8.2) | 2400 (0.1) |
| >50 yr | 606,631 | 329,270 (54.3) | 257,075 (42.4) | 293,619 (48.4) | 255,678 (42.1) | 35,651 (5.9) | 1397 (0.2) |
| Not defined | n/a | 105 | 44 | 97 | 44 | 8 | 0 |
| **Sedibeng** | | | | | | | |
| **Overall** | **771,158** | **218,705 (28.4)** | **109,121 (14.2)** | **144,983 (18.8)** | **108,287 (14.0)** | **73,722 (9.6)** | **834 (0.1)** |
| 12–18 yr | 101,461 | 7838 (7.7) | 1362 (1.3) | 6101 (6.0) | 1362 (1.3) | 1737 (1.7) | 0 |
| >18 to 50 yr | 475,958 | 119,966 (25.2) | 46,646 (9.8) | 66,441 (14.0) | 46,165 (9.7) | 53,525 (11.2) | 481 (0.1) |
| >50 yr | 193,738 | 90,881 (46.9) | 61,104 (31.5) | 72,424 (37.4) | 60,751 (31.4) | 18,457 (9.5) | 353 (0.2) |
| Not defined | n/a | 20 | 9 | 17 | 9 | 3 | 0 |
| **City of Tshwane** | | | | | | | |
| **Overall** | **2,933,611** | **1,029,116 (35.1)** | **531,597 (18.1)** | **675,860 (23.0)** | **524,529 (17.9)** | **353,256 (12.0)** | **7068 (0.2)** |
| 12–18 yr | 356,800 | 31,733 (8.9) | 7533 (2.1) | 26,069 (7.3) | 7519 (2.1) | 5664 (1.6) | 14 (<0.1) |
| >18 to 50 yr | 1,996,039 | 621,768 (31.2) | 254,539 (12.8) | 356,047 (17.8) | 250,172 (12.5) | 265,721 (13.1) | 4367 (0.2) |
| >50 yr | 580,771 | 375,470 (64.7) | 269,450 (46.4) | 293,632 (50.6) | 266,763 (45.9) | 81,838 (14.1) | 2687 (0.5) |
| Not defined | n/a | 145 | 75 | 112 | 75 | 33 | 0 |
| **West Rand** | | | | | | | |
| **Overall** | **752,898** | **298,698 (39.7)** | **158,946 (21.1)** | **221,055 (29.4)** | **158,547 (21.1)** | **77,643 (10.3)** | **399 (0.1)** |
| 12–18 yr | 94,744 | 12,255 (12.9) | 1654 (1.7) | 9940 (10.5) | 1654 (1.7) | 2315 (2.4) | 0 |
| >18 to 50 yr | 489,114 | 189,703 (38.8) | 86,313 (17.6) | 129,543 (26.5) | 86,061 (17.6) | 60,160 (12.3) | 252 (0.1) |
| >50 yr | 169,039 | 96,721 (57.2) | 70,972 (42.0) | 81,555 (48.2) | 70,825 (41.9) | 15,166 (9.0) | 147 (0.1) |
| Not defined | n/a | 19 | 7 | 17 | 7 | 2 | 0 |

*The current recommended dosing schedule for BNT162b2 in South Africa is 2 doses for individuals aged >18 years and 1 dose for children aged 12–18 years. †The current recommended dosing schedule of Ad26.COV2.S in South Africa is 1 dose for all age groups. A booster dose is available to healthcare workers through the Sisonke2 study. n/a, not applicable

**Table S4: Seroprevalence of SARS-CoV-2 anti-spike (anti-S) or anti-nucleocapsid (anti-N) immunoglobulin G (IgG) Gauteng Province across the Districts and sub-Districts irrespective of Covid-19 vaccination status.**

| District / sub-district | Population – no.* | Participants in survey | Anti-S or anti-N – no. (%) [95% CI] | Anti-S IgG reactive only – no.(%) [95% CI] | Overall anti-N IgG reactive – no. (%) [95% CI] | Overall anti-S IgG reactive – no. (%) [95%] CI) |
|---|---|---|---|---|---|---|
| **Gauteng Province** | **15,176,113** | **7010** | **5125 (73.1) [72.0–74.1]** | **2275 (32.5) [31.4–33.5]** | **2850 (40.7) [39.5–41.8]** | **4906 (70.0) [68.9–71.0]** |
| **Johannesburg District** | **5,606,238** | **2468** | **1881 (76.2) [74.5–77.8]** | **794 (32.2) [30.3–34.0]** | **1087 (44.0) [42.1–46.0]** | **1792 (72.6) [70.8–74.3]** |
| Johannesburg A | 779,519 | 333 | 246 (73.9) [68.9–78.3] | 111 (33.3) [28.5–38.6] | 135 (40.5) [35.4–45.9] | 231 (69.4) [64.2–74.1] |
| Johannesburg B | 435,241 | 197 | 169 (85.8) [80.2–90.0] | 55 (27.9) [22.1–34.6] | 114 (57.9) [50.9–64.6] | 152 (77.2) [70.8–82.5] |
| Johannesburg C | 799,980 | 444 | 363 (81.8) [77.9–85.1] | 139 (31.3) [27.2–37.2] | 224 (50.5) [45.8–43.5] | 356 (80.2) [76.2–83.6] |
| Johannesburg D | 1,396,243 | 646 | 472 (73.1) [69.5–76.3] | 216 (33.4) [29.9–37.2] | 256 (39.6) [35.9–43.5] | 450 (69.7) [66.0–73.1] |
| Johannesburg E | 601,433 | 161 | 117 (72.7) [65.3–79.0] | 60 (37.3) [30.1–45.0] | 57 (35.4) [28.4–43.1] | 113 (70.2) [62.7–76.7] |
| Johannesburg F | 751,484 | 243 | 185 (76.1) [70.4–81.1] | 80 (32.9) [27.3–39.1] | 105 (43.2) [37.1–49.5] | 172 (70.8) [64.8–76.2] |
| Johannesburg G | 842,339 | 444 | 329 (74.1) [69.7–77.8] | 133 (30.0) [25.8–34.3] | 196 (44.1) [39.5–48.7] | 318 (71.6) [67.1–75.5] |
| **Ekurhuleni District** | **3,825,650** | **1861** | **1382 (74.3) [72.2–76.2]** | **597 (32.1) [30.0–34.2]** | **785 (42.2) [40.0–44.4]** | **1305 (70.1) [68.0–72.2]** |
| Ekurhuleni E1 | 626 517 | 353 | 242 (68.6) [63.5–73.2] | 124 (35.1) [30.3–40.3] | 118 (33.4) [28.7–38.5] | 229 (64.9) [59.7–69.7] |
| Ekurhuleni E2 | 455 262 | 252 | 190 (75.4) [69.7–80.3] | 81 (32.1) [26.7–38.2] | 109 (43.3) [37.3–49.4] | 181 (71.8) [66.0–77.0] |
| Ekurhuleni N1 | 708 290 | 358 | 244 (68.2) [63.1–72.8] | 124 (34.6) [29.9–39.7] | 120 (33.5) [28.8–38.6] | 232 (64.8) [59.7–69.6] |
| Ekurhuleni N2 | 697 175 | 258 | 206 (79.8) [74.5–84.3] | 63 (24.4) [19.6–30.0] | 143 (55.4) [49.3–61.4] | 177 (68.6) [62.7–74.0] |
| Ekurhuleni S1 | 673 758 | 210 | 172 (81.9) [76.1–86.5] | 76 (36.2) [30.0–42.9] | 96 (45.7) [39.1–52.5] | 169 (80.5) [74.6–85.3] |
| Ekurhuleni S2 | 664 648 | 430 | 328 (76.3) [72.0–80.1] | 129 (30.0) [25.9–34.5] | 199 (46.3) [41.6–51.0] | 317 (73.7) [69.4–77.7] |
| **Sedibeng District** | **1,084,503** | **564** | **397 (70.4)** | **188 (33.3)** | **209 (37.1)** | **387 (68.6)** |

| | | | | | | |
|---|---|---|---|---|---|---|
| | | | **[66.5–74.0]** | **[29.6–37.3]** | **[33.2–41.1]** | **[64.7–72.3]** |
| Lesedi | 127 419 | 408 | 292 (71.6) [67.0–75.7] | 150 (36.8) [32.2–41.6] | 142 (34.8) [30.3–39.6] | 286 (70.1) [65.5–74.3] |
| Midvaal | 126 285 | 104 | 65 (62.5) [52.8–71.3] | 26 (25.0) [17.6–34.2] | 39 (37.5) [28.7–47.2] | 63 (60.6) [50.9–69.5] |
| Emfuleni | 830 798 | 52 | 40 (76.9) [63.6–86.4] | 12 (23.1) [13.6–36.4] | 28 (53.8) [40.3–66.8] | 38 (73.1) [59.5–83.4] |
| **City of Tshwane District** | **3,709,635** | **1464** | **976 (66.7) [54.2–69.0]** | **471 (32.2) [29.8–34.6]** | **505 (34.5) [32.1–36.9]** | **939 (64.1) [61.6–66.5]** |
| Tshwane 1 | 1 032 885 | 471 | 298 (63.3) [58.8–67.5] | 145 (30.8) [26.8–35.1] | 153 (32.5) [28.4–36.8] | 287 (60.9) [56.4–65.2] |
| Tshwane 2 | 436 950 | 175 | 103 (58.9) [51.4–65.9] | 57 (32.6) [26.0–39.9] | 46 (26.3) [20.3–33.3] | 99 (56.6) [49.1–63.7] |
| Tshwane 3 | 730 788 | 229 | 177 (77.3) [71.4–82.3] | 72 (31.4) [25.8–37.7] | 105 (45.9) [39.5–52.3] | 167 (72.9) [66.8–78.3] |
| Tshwane 4 | 482 448 | 78 | 46 (59.0) [47.8–69.3] | 19 (24.4) [16.1–35.1] | 27 (34.6) [24.9–45.8] | 44 (56.4) [45.3–66.9] |
| Tshwane 5 | 119 190 | 204 | 129 (63.2) [56.4–69.6] | 72 (35.3) [29.0–42.1] | 57 (27.9) [22.2–34.5] | 126 (61.8) [54.9–68.2] |
| Tshwane 6 | 768 446 | 245 | 175 (71.4) [65.5–76.7] | 89 (36.3) [30.5–42.5] | 86 (35.1) [29.4–41.3] | 169 (69.0) [62.9–74.5] |
| Tshwane 7 | 138 928 | 62 | 48 (77.4) [64.2–85.1] | 17 (27.4) [17.5–39.2] | 31 (50.0) [37.2–61.4] | 47 (75.8) [62.5–83.8] |
| **West Rand District** | **950,088** | **653** | **489 (74.9) [71.4–78.1]** | **225 (34.5) [30.9–38.2]** | **264 (40.4) [36.7–44.2]** | **483 (74.0) [70.5–77.2]** |
| Mogale City | 435 254 | 149 | 95 (63.8) [55.7–71.1] | 51 (34.2) [27.1–42.2] | 44 (29.5) [22.8–37.3] | 95 (63.8) [55.7–71.1] |
| Rand West City | 300 960 | 261 | 208 (79.7) [74.4–84.1] | 92 (35.2) [29.7–41.2] | 116 (44.4) [38.5–50.5] | 207 (79.3) [74.0–83.8] |
| Merafong City | 213 874 | 243 | 196 (76.5) [70.8–81.4] | 82 (33.7) [28.1–39.9] | 104 (42.8) [36.7–49.1] | 181 (74.5) [68.6–79.6] |

CI, confidence interval
Seroprevalence was calculated as the number of individuals seropositive for anti-S or anti-N IgG divided by the total number of individuals sampled. Confidence intervals have not been adjusted for multiplicity and should not be used for inference.

**Table S5. Number and incidence per 100,000 population of Covid-19 cases, in Gauteng Province by Covid-19 wave, stratified by age and by gender.**

| | Wave 1* | Wave 2* | Wave 3* | Wave 4* | TOTAL |
|---|---|---|---|---|---|
| **Dominant variant** | Wild type | Beta | Delta | Omicron | |
| **Period of wave** | Mar 7–Nov 13, 2020 | Nov 14, 2020–Mar 30, 2021 | Mar 31–Oct 25, 2021 | Oct 26, 2021–Jan 12, 2022 | Mar 7, 2020–Jan 12, 2022 |
| **Age group: 0–4 years** | | | | | |
| **Cases in wave – no.†** | 2761 | 2005 | 6709 | 4724 | 16,199 |
| **Case rate per 100,000 population** | 212 | 154 | 514 | 362 | 1241 |
| **Proportion of total cumulative cases, %** | 17.0 | 12.4 | 41.4 | 29.2 | 100 |
| **Age group: 5–19 years** | | | | | |
| **Cases in wave – no.†** | 14,798 | 12,892 | 53,636 | 26,067 | 107,393 |
| **Case rate per 100,000 population** | 435 | 379 | 1575 | 766 | 3154 |
| **Proportion of total cumulative cases, %** | 13.8 | 12.0 | 49.9 | 24.3 | 100 |
| **Age group: 20–49 years** | | | | | |
| **Cases in wave – no.†** | 148,240 | 112,919 | 300,032 | 147,270 | 708,461 |
| **Case rate per 100,000 population** | 1818 | 1385 | 3680 | 1806 | 8690 |
| **Proportion of total cumulative cases, %** | 20.9 | 15.9 | 42.3 | 20.8 | 100 |
| **Age group: 50–59 years** | | | | | |
| **Cases in wave – no.†** | 35,434 | 27,584 | 81,120 | 25,408 | 169,546 |
| **Case rate per 100,000 population** | 2695 | 2098 | 6169 | 1932 | 12,894 |
| **Proportion of total cumulative cases, %** | 20.9 | 16.3 | 47.8 | 15.0 | 100 |
| **Age group: >60 years** | | | | | |
| **Cases in wave – no.†** | 30,896 | 27,164 | 70,142 | 23,462 | 151,664 |
| **Case rate per 100,000 population** | 2358 | 2073 | 5353 | 1791 | 11,576 |
| **Proportion of total cumulative cases, %** | 20.4 | 17.9 | 46.2 | 15.5 | 100 |
| **Sex: male‡** | | | | | |
| **Cases in wave – no.†** | 98,223 | 80,612 | 229,407 | 99,837 | 508,079 |
| **Case rate per 100,000 population** | 1267 | 1040 | 2958 | 1287 | 6552 |
| **Proportion of total cumulative cases, %** | 19.3 | 15.9 | 45.2 | 19.6 | 100 |
| **Sex: female‡** | | | | | |
| **Cases in wave – no.†** | 132,425 | 100,661 | 278,826 | 125,338 | 637,250 |
| **Case rate per 100,000 population** | 1712 | 1302 | 3605 | 1621 | 8240 |
| **Proportion of total cumulative cases, %** | 20.8 | 15.8 | 43.8 | 19.7 | 100.0 |

*All data are from the National Institute for Communicable Diseases daily databases. The Omicron-dominant fourth case wave is at its tail-end but has not yet fully subsided. Totals, incidence, and proportions of cases are anticipated to continue to increase somewhat over the next few weeks until the wave has fully subsided. †Changes in testing rates, particularly the lower rates during Wave 1 due to constraints in laboratory capacity and prioritization of testing for hospitalized individuals, prevent direct comparisons, especially in terms of case numbers during the first wave in relation to the subsequent waves. Cases include asymptomatic and symptomatic individuals. Cumulative reported cases were sourced from the National Department of Health.[12] ‡Sex was not recorded for 7895 Covid-19 cases, who are excluded from analyses by sex.

**Table S6. Number and incidence per 100,000 population of hospitalizations in Gauteng Province by Covid-19 wave, stratified by age and by gender.**

| | Wave 1* | Wave 2* | Wave 3* | Wave 4* | TOTAL |
|---|---|---|---|---|---|
| **Dominant variant** | Wild type | Beta | Delta | Omicron | |
| **Period of wave** | Mar 7–Nov 13, 2020 | Nov 14, 2020–Mar 30, 2021 | Mar 31–Oct 25, 2021 | Oct 26, 2021–Jan 12, 2022 | Mar 7, 2020–Jan 12, 2022 |
| **Age group: 0–4 years** | | | | | |
| **Hospitalisations in wave – no.†** | 523 | 724 | 1377 | 1288 | 3912 |
| **Hospitalisation rate per 100,000 population** | 40 | 55 | 106 | 99 | 300 |
| **Proportion of total cumulative hospitalisations, %** | 13.4 | 18.5 | 35.2 | 32.9 | 100 |
| **Age group: 5–19 years** | | | | | |
| **Hospitalisations in wave – no.†** | 745 | 683 | 1551 | 1118 | 4097 |
| **Hospitalisation rate per 100,000 population** | 22 | 20 | 46 | 33 | 120 |
| **Proportion of total cumulative hospitalisations, %** | 18.2 | 16.7 | 37.9 | 27.3 | 100 |
| **Age group: 20–49 years** | | | | | |
| **Hospitalisations in wave – no.†** | 13,882 | 11,914 | 21,028 | 7650 | 54,474 |
| **Hospitalisation rate per 100,000 population** | 170 | 146 | 258 | 94 | 668 |
| **Proportion of total cumulative hospitalisations, %** | 25.5 | 21.9 | 38.6 | 14.0 | 100 |
| **Age group: 50–59 years** | | | | | |
| **Hospitalisations in wave – no.†** | 6755 | 6828 | 12,888 | 1673 | 28,144 |
| **Hospitalisation rate per 100,000 population** | 514 | 519 | 980 | 127 | 2140 |
| **Proportion of total cumulative hospitalisations, %** | 24.0 | 24.3 | 45.8 | 5.9 | 100 |
| **Age group: >60 years** | | | | | |
| **Hospitalisations in wave – no.†** | 11,712 | 11,318 | 23,615 | 4159 | 50,804 |
| **Hospitalisation rate per 100,000 population** | 894 | 864 | 1802 | 317 | 3878 |
| **Proportion of total cumulative hospitalisations, %** | 23.1 | 22.3 | 46.5 | 8.2 | 100 |
| **Sex: male‡** | | | | | |
| **Hospitalisations in wave – no.†** | 15,237 | 14,590 | 29,255 | 6663 | 65,745 |
| **Hospitalisation rate per 100,000 population** | 196 | 188 | 377 | 86 | 848 |
| **Proportion of total cumulative hospitalisations, %** | 23.2 | 22.2 | 44.5 | 10.1 | 100 |
| **Sex: female‡** | | | | | |
| **Hospitalisations in wave – no.†** | 18,323 | 16,850 | 31,155 | 9198 | 75,526 |
| **Hospitalisation rate per 100,000 population** | 237 | 218 | 403 | 119 | 977 |
| **Proportion of total cumulative hospitalisations, %** | 24.3 | 22.3 | 41.3 | 12.2 | 100 |

*All data are from DATCOV, hosted by the National Institute for Communicable Disease,[7] as described previously.[9] The system was developed during the course of the first wave, with gradual onboarding of facilities; hence, these data could underestimate hospitalized cases in the first wave relative to subsequent waves. The hospitalized cases include individuals with Covid-19, as well as coincidental infections identified as part of routine testing for SARS-CoV-2 of individuals admitted to the facilities to assist in triaging of patients in the hospital. The Omicron-dominant fourth case wave is at its tail-end but has not yet fully subsided. Totals, incidence, and proportions of hospitalizations are anticipated to continue to increase somewhat over the next few weeks until the wave has fully subsided. †Changes in testing rates, particularly the lower rates during Wave 1 due to constraints in laboratory capacity and prioritization of testing for hospitalized individuals, prevent direct comparisons, especially in terms of case numbers during the first wave in relation to the subsequent waves. ‡Sex was not recorded for 162 hospitalizations, who are excluded from analyses by sex.

**Table S7. Number and incidence per 100,000 population of recorded deaths in Gauteng Province by Covid-19 wave, stratified by age and by gender.**

| | Wave 1* | Wave 2* | Wave 3* | Wave 4* | TOTAL |
|---|---|---|---|---|---|
| **Dominant variant** | Wild type | Beta | Delta | Omicron | |
| **Period of wave** | Mar 7–Nov 13, 2020 | Nov 14, 2020–Mar 30, 2021 | Mar 31–Oct 25, 2021 | Oct 26, 2021–Jan 6, 2022 | Mar 7, 2020–Jan 6, 2022 |
| **Age group: 0–4 years** | | | | | |
| **Recorded deaths in wave – no.** | 36 | 22 | 49 | 19 | 126 |
| **Recorded death rate per 100,000 population** | 3 | 2 | 4 | 1 | 10 |
| **Proportion of total cumulative recorded deaths, %** | 28.6 | 17.5 | 38.9 | 15.1 | 100 |
| **Age group: 5–19 years** | | | | | |
| **Recorded deaths in wave – no.** | 24 | 15 | 51 | 22 | 112 |
| **Recorded death rate per 100,000 population** | 1 | 0 | 1 | 1 | 3 |
| **Proportion of total cumulative recorded deaths, %** | 21.4 | 13.4 | 45.5 | 19.6 | 100 |
| **Age group: 20–49 years** | | | | | |
| **Recorded deaths in wave – no.** | 1184 | 1309 | 2566 | 299 | 5358 |
| **Recorded death rate per 100,000 population** | 15 | 16 | 31 | 4 | 66 |
| **Proportion of total cumulative recorded deaths, %** | 22.1 | 24.4 | 47.9 | 5.6 | 100 |
| **Age group: 50–59 years** | | | | | |
| **Recorded deaths in wave – no.** | 1195 | 1465 | 3144 | 146 | 5950 |
| **Recorded death rate per 100,000 population** | 91 | 111 | 239 | 11 | 452 |
| **Proportion of total cumulative recorded deaths, %** | 20.1 | 24.6 | 52.8 | 2.5 | 100 |
| **Age group: >60 years** | | | | | |
| **Recorded deaths in wave – no.** | 3730 | 4058 | 8873 | 692 | 17,353 |
| **Recorded death rate per 100,000 population** | 285 | 310 | 677 | 53 | 1324 |
| **Proportion of total cumulative recorded deaths, %** | 21.5 | 23.4 | 51.1 | 4.0 | 100 |
| **Sex: male‡** | | | | | |
| **Recorded deaths in wave – no.** | 3245 | 3317 | 7806 | 601 | 14,969 |
| **Recorded death rate per 100,000 population** | 42 | 43 | 101 | 8 | 193 |
| **Proportion of total cumulative recorded deaths, %** | 21.7 | 22.2 | 52.1 | 4.0 | 100 |
| **Sex: female†** | | | | | |
| **Recorded deaths in wave – no.** | 2919 | 3551 | 6864 | 573 | 13,907 |
| **Recorded death rate per 100,000 population** | 38 | 46 | 89 | 7 | 180 |
| **Proportion of total cumulative recorded deaths, %** | 21.0 | 25.5 | 49.4 | 4.1 | 100 |

*All data are from the National Department of Health.[12] The Omicron-dominant fourth case wave is at its tail-end but has not yet fully subsided. Totals, incidence, and proportions of deaths are anticipated to continue to increase somewhat over the next few weeks until the waves has fully subsided. †Sex was not recorded for 22 recorded deaths, who are excluded from analyses by sex.
